# Prolonged T-cell activation and long COVID symptoms independently associate with severe disease at 3 months in a UK cohort of hospitalized COVID-19 patients

**DOI:** 10.1101/2022.11.25.22282759

**Authors:** Marianna Santopaolo, Michaela Gregorova, Fergus Hamilton, David Arnold, Anna Long, Aurora Lacey, Elizabeth Oliver, Alice Halliday, Holly Baum, Kristy Hamilton, Rachel Milligan, Olivia Pearce, Lea Knezevic, Begonia Morales Aza, Alice Milne, Emily Milodowski, Rajeka Lazarus, Anu Goenka, Adam Finn, Nick Maskell, Andrew Davidson, Kathleen Gillespie, Linda Wooldridge, Laura Rivino

## Abstract

COVID-19 causes immune perturbations which may persist long-term, and patients frequently report ongoing symptoms for months after recovery. We assessed the extent and nature of immune activation at 3 months post hospital admission in patients with mild, moderate or severe COVID-19 and investigated whether immune activation associates with disease severity and long COVID. Patients with severe disease displayed persistent activation of CD4^+^ and CD8^+^ T-cells, based on expression of HLA-DR, CD38, Ki67 and granzyme B, but they lacked activation of other immune subsets. Elevated plasma levels of IL-4, IL-7, IL-17 and TNF-α were present in patients with severe compared to mild and/or moderate disease. Plasma from severe patients caused T-cells from healthy donors to upregulate IL-15Rα, suggesting that factors in the plasma of severe patients may increase T-cell responsiveness to IL-15-driven ‘bystander” activation, which may drive persistent T-cell activation after severe COVID-19. Patients with severe disease reported a higher number of long COVID symptoms which correlated with the frequency of two subsets of activated CD4^+^ and CD8^+^ T cells (CD4^+^ T-cell population 2 and CD8^+^ T-cell population 4; FDR p<0.05), however these associations were lost after adjusting for age, sex and disease severity. Our data suggests that persistent immune activation and long COVID correlate independently with severe disease.

## Introduction

Infection with severe acute respiratory syndrome coronavirus 2 (SARS-CoV-2), the causative agent of Coronavirus Disease-19 (COVID-19), can be asymptomatic or lead to a broad spectrum of disease manifestations from mild to severe disease and death. There is evidence showing that acute COVID-19 causes a profound activation of innate and adaptive immune cells, and distinct immunological signatures are shown to correlate with disease outcomes^1^. Viral control and mild disease associate with the rapid generation of T-cells and antibodies targeting SARS-CoV-2^2, 3^, while severe disease is characterized by immune cell hyperactivation and a “cytokine storm”^4–6^. Commonly observed biomarkers of severe COVID-19 include lymphopenia, increased neutrophil to T-cell ratio and elevated levels of pro-inflammatory cytokines/mediators such as IL-6, IL-10, IL-17, MCP-1, IP-10, CRP, IL-1Rα and IL-1β. In a smaller proportion of individuals severe COVID-19 may be driven by pre-existing anti-IFN-γ autoantibodies^7–10^. Other distinctive features of severe disease include a dysregulation in the myeloid compartment consisting of increased circulating immature/dysfunctional neutrophils and immature Ki67^+^ COX-2^low^ monocytes^11–14^, while T-cell activation and proliferation appears to be heterogenous in severe patients^1, 5^.

The kinetics of immune recovery following the changes occurring during acute COVID-19 are complex and not fully understood. Longitudinal studies show that immune abnormalities and inflammation may persist after severe COVID-19, with highly activated myeloid cells, pro-inflammatory cytokines and persistently activated T-cells detected 8-12 months after COVID-19^15,16,17^.

The potential impact of prolonged immune activation post recovery and whether it may underlie the symptoms of post-acute sequelae of SARS-CoV-2 infection (PASC) or long COVID - a syndrome characterized by long-lasting symptoms affecting multiple organs - remains unclear. Long COVID is observed in 8-21% of individuals following mild to severe COVID-19, although a higher prevalence of symptoms is reported in patients requiring ICU admission and/or mechanical ventilation^18–20^. Several studies have reported a correlation between long COVID symptoms and a variety of immune profiles including a more rapid decline of SARS-CoV-2 memory CD8^+^ T-cells and lower levels of cytotoxic SARS-CoV-2 N-specific CD8^+^ T-cells^21^, increased levels of cytotoxic CD8^+^ T-cells and elevated plasma levels of type I cytokines, IL-1β and IL-6^22, 23^. Another longitudinal study conducted in 69 patients found long COVID to associate with an altered immune blood cell transcriptome which included significant changes in genes involved in cell cycle, but not with the phenotypic profiles of these cells^24^. A large UK cohort study (PHOSP-COVID) of hospitalized COVID-19 patients identified female sex, obesity and invasive mechanical ventilation as factors associated with a lower likelihood of full recovery, with an overall recovery rate of 28.9% at both 5 and 12 months across all disease severities^25^. Similarly, other cohort studies have identified associations between poor recovery from COVID-19 and demographic factors such as sex and age, but also highlight a possible role of immune dysregulation that occurs in older age, obesity and asthma, which warrants further investigation^26,27,28^. Other hypotheses proposed to explain the mechanisms underlying long COVID include unresolved lung tissue damage, the persistence of SARS-CoV-2 infection, the reactivation of latent viruses such as human cytomegalovirus (CMV) and Epstein-Barr virus (EBV) and autoimmunity^29–32^. Due to the limited effectiveness of COVID-19 vaccines in protecting from re-infection, SARS-CoV-2 continues to circulate within our populations and millions of people world-wide are at risk of experiencing long-term health complications from COVID-19. Therefore, the burden of long COVID threatens to increase and further compromise our post-pandemic economic recovery. A better understanding of the mechanisms underlying long COVID and the design/repurposing of drugs to treat or prevent this syndrome is urgently needed.

In this study we investigated whether patients who had COVID-19, 3 months post hospitalization display persistent immune activation and ongoing inflammation and if there are potential links between the observed immune profiles, COVID-19 disease severity and long COVID symptoms. We also investigated whether persistent immune activation and disease severity have an impact on the capacity of patients to generate and maintain a memory T-cell response and antibodies to SARS-CoV-2. Our immunological analysis of 154 samples from 63 hospitalized patients recovering from mild, moderate or severe disease showed increased levels of activated CD4^+^ and CD8^+^ T-cells and increased plasma levels of T-cell-related cytokines (IL-4, IL-7, IL-17 and TNF-α) at 3 months in severe compared to moderate/mild patients. The SARS-CoV-2-specific memory T-cell response to spike, membrane and nucleocapsid was robust and qualitatively similar across the disease severities, however the magnitude of the CD4^+^ and CD8^+^ T-cell and antibody response was higher in patients with moderate compared to mild disease, as were the plasma levels of IFN-γ, a key cytokine for anti-viral immunity. Long COVID symptoms at 3 months were reported in 80% of patients across all disease severities but were more frequent in patients with severe disease. Poisson regression analysis showed a significant association between ongoing symptoms and the frequency of a subset of moderately activated CD4^+^ T and non-cytotoxic HLA-DR^+^ CD8^+^ T-cells in unadjusted analyses, but these associations did not meet the criteria for statistical significance after adjusting for age, sex and severity grades. Our results demonstrate the presence of immune perturbations in peripheral blood CD4^+^ and CD8^+^ T-cell populations in severe COVID-19 patients 3 months after hospitalization with no direct association between long COVID symptoms and immune activation/pro-inflammatory cytokines, for the markers that were measured.

## Results

### DISCOVER patient cohort

To investigate immune profiles in convalescent COVID-19 patients we obtained peripheral blood mononuclear cells (PBMCs) and plasma from 63 patients enrolled in the DISCOVER study at 3 months post admission for COVID-19, and where possible matched plasma samples at 8 and 12 months. The demographics and clinical characteristics of the 63 patients included in this study are shown in **Table 1**. Patients were stratified into mild, moderate and severe based on the type of respiratory support they required during the acute illness, as follows: mild patients did not require supplementary oxygen or intensive care; moderate patients required supplementary oxygen during admission and severe patients required invasive mechanical ventilation, non-invasive ventilation and/or admission to the intensive treatment unit (ITU). Of the 63 patients included in this analysis 17, 32 and 14 recovered from mild, moderate and severe disease, respectively. Overall, a higher proportion of patients were male (40/63) and the median age (± SD) of patients was 53 ± 14.5, 58 ± 12.6 and 61.5 ± 10 years for mild, moderate and severe patients, respectively. Patient ethnicity was predominantly Caucasian with an Asian/Black minority. Body Mass Index (BMI) was largely within the unhealthy range across all disease severities (overweight, obese and extremely obese) with 7.6%, 15.6% and 14.3% of patients with respectively mild, moderate and severe disease displaying healthy BMI. The comorbidities observed in these 63 patients were consistent with those known to be associated with higher risks for COVID-19 hospitalization and included heart disease, diabetes (predominantly type-2), hypertension and chronic lung disease. Overall, the percentage of patients with comorbidities tended to increase progressively with disease severity (mild: 52.9%; moderate: 56.2 % and severe: 85.7%). The duration of hospital admission ranged from 3.3 ± 1.99 to 7.8 ± 5.16 and 12 ± 6.67 days (± SD) in mild, moderate and severe patients, respectively. None of the patients had received COVID-19 vaccination at the time of blood collection. Overall, at 3 months post admission patients’ blood lymphocyte and neutrophil counts, CRP and albumin levels - which were mostly perturbated during the acute illness - had normalized to levels that remained similar at 8 months, suggesting a resolution of the peak inflammatory events occurring during the acute illness (**Supplementary Figure 1**).

**Table 1.** Details of the patients included in this

| Disease severity (n) |  | Mild<br>(n = 17) | Moderate<br>(n = 32) | Severe<br>(n = 14) |
| --- | --- | --- | --- | --- |
| Age (median, SD) |  | 53 ± 14.5 | 58 ± 12.6 | 61.5 ±10 |
| Sex, %(n) | Female | 35.3 (6) | 31.2 (10) | 50 (7) |
|  | Male | 64.7 (11) | 68.5 (22) | 50 (7) |
| Ethnicity, % (n) | Caucasian | 83.3% (14) | 78.1% (25) | 87.7% (12) |
|  | Asian | 11.7% (2) | 15.6% (5) | 14.3% (2) |
|  | Black | 5.9% (1) | 6.3% (2) | 0 (0) |
| BMI, kg/m2, %<br>(average, SD) | Healthy | 7.6% (22.3±0.57) | 15.6% (23±1.73) | 14.3% (23±0) |
|  | Overweight | 35.3% (26.6±1.5) | 28.1% (27±1.39) | 28.6% (28.75±0.5) |
|  | Obese | 35.3%(32.8±1.47) | 46.9% (32.7±2.46) | 28.6% (35±2.1) |
|  | Extremely Obese | 11.8% (61±28.2) | 9.38% (46.3±6.02) | 28.6% (46±7.34) |
| Comorbidity % (n) | None | 47.1% (8) | 43.75% (14) | 14.3% (2) |
|  | Heart disease | 23.5% (4) | 15.6% (5) | 14.3% (2) |
|  | T1DM | 0 (0) | 3.1% (1) | 7.14% (1) |
|  | T2DM | 5.88% (1) | 12.5% (4) | 14.3% (2) |
|  | Hypertension | 17.65 (3) | 18.75% (6) | 50% (7) |
|  | Chronic lung disease | 5.88% (1) | 21.88% (7) | 50% (7) |
|  | Kidney disease | 5.88 (1) | 9.37% (3) | 14.3% (2) |
|  | Mental health | 0 (0) | 6.25% (2) | 14.3% (2) |
|  | Cancer | 5.88% (1) | 3.1% (1) | 7.14 (1) |
|  | Asthma | 5.88% (1) | 0 (0) | 14.3% (2) |
|  | Total Obesity | 47.1% (8) | 53.1% (17) | 57.1% (8) |
|  | Other | 47.1% (8) | 28.1% (9) | 50% (7) |
| Hospital stay (average days, SD) |  | 3.3±1.99 | 7.8±5.16 | 12±6.67 |
| Ongoing symptoms<br>at 3 months (n, %) | None | 3(17.65%) | 8(25%) | 2(14.3%) |
|  | Dyspnoea | 7(41.2%) | 15(47%) | 9(64.3%) |
|  | Excessive fatigue | 5(29.4%) | 12(37.5%) | 9(64.3%) |
|  | Muscle weakness | 3(17.65) | 7(22%) | 5(35.7%) |
|  | Sleeping difficulties | 3(17.65) | 6(18.75%) | 8 (57%)) |
|  | Psychiatric | 2(11.8%) | 8(25%) | 6(42.9%) |
|  | Anosmia | 2(11.8%) | 4(12.5%) | 3 (21.4%) |
|  | Chest pain | 2(11.8%) | 6(18.75%) | 2(14.3%) |
|  | Cough | 2(11.8%) | 4(12.5%) | 1 (7.14%) |
|  | Other | 5(29.4%) | 4(12.5%) | 3 (21.4%) |

n= number of patients; SD=standard deviation.

### Increased CD4^+^ and CD8^+^ T-cell activation in severe patients at 3 months

To investigate the presence of ongoing immune activation following recovery from COVID-19, we assessed the phenotype and activation/proliferation profiles of conventional CD4^+^ and CD8^+^ T-cells, TCR-γδ T, NK, B-cells and monocytes by flow cytometry in PBMC samples from 63 patients at 3 months post admission. T and NK cells were assessed for expression of markers of activation, differentiation, proliferation and cytotoxicity (HLA-DR, CD38, CD69, CCR7, CD45RA, CXCR3, Ki67, granzyme B, CD56) after gating on single live cells, CD3^+^ CD4^+^ or CD3^+^ CD8^+^ T-cells and CD3^-^ CD56^bright^ CD16^+/-^ or CD56^dim^ CD16^+^ NK cell populations, respectively (gating strategies in **Supplementary Figure 2**). Data were analysed by FlowJo using a manual gating strategy as well as the dimensionality reduction algorithm Uniform Manifold Approximation and Projection for Dimension Reduction (UMAP) and cluster analysis using FlowSOM.

The frequencies and absolute numbers of CD4^+^ T-cells were similar in patients across disease severities (**Figure 1 A, B**). However, we observed an increased frequency of CD4^+^ T-cells expressing the peripheral homing receptor CXCR3 in severe compared to moderate patients and an increased frequency of CD4^+^ T-cells co-expressing the proliferation/activation markers Ki67 and CD38 in severe compared to mild patients (**Figure 1** **C, D**). The overall differentiation status of CD4^+^ T-cells, defined by the coordinated expression of CCR7 and CD45RA was similar in mild, moderate and severe patients, with high proportions of naïve (CCR7^+^ CD45RA^+^) and T central memory cells (CCR7^+^ CD45RA^-^, T_CM_) followed by detectable levels of T effector memory cells (CCR7^-^ CD45RA^-^, TEM) and low frequencies of T effector memory re-expressing RA cells (CCR7^-^ CD45RA^+^, T_EMRA_) (**Figure 1** **E**). The most abundant naïve and T_CM_ CD4^+^ T-cell subsets contained significantly higher levels of cells co-expressing markers of activation, proliferation and cytotoxic potential (HLA-DR^+^ CD38^+^, HLA-DR^+^ Ki67^+^, CD38^+^ granzyme B^+^) in severe compared to moderate/mild patients (**Figure 1** **F-K**). An unsupervised analysis by UMAP and FlowSOM revealed the presence of 15 distinct clusters of cells characterized by a unique, coordinated expression of the analyzed markers (**Figure 1** **L**), shown as Mean Fluorescence Intensity (MFI) in the heatmap (**Figure 1** **M**). These analyses revealed significantly higher frequencies in severe compared to moderate patients of clusters of cells expressing HLA-DR, CD38 and Ki67 (populations 8 and 13) as well as of cells that express CCR7 but lack expression of all other markers analyzed, and which resemble undifferentiated, resting CD4^+^ T-cells (population 2) (**Figure 1** **N**). The latter population accounted for a large proportion of CD4^+^ T-cells (up to 60% of cells in severe patients). A population characterized by low levels of expression of all markers analyzed is decreased in severe compared to moderate patients (population 1, **Figure 1 N**). In summary, both manual gating and unsupervised analysis reveal the presence of higher frequencies of CD4^+^ T-cell populations expressing markers of activation, proliferation and peripheral tissue homing in patients who recovered from severe versus mild and/or moderate disease.

**Figure 1.**
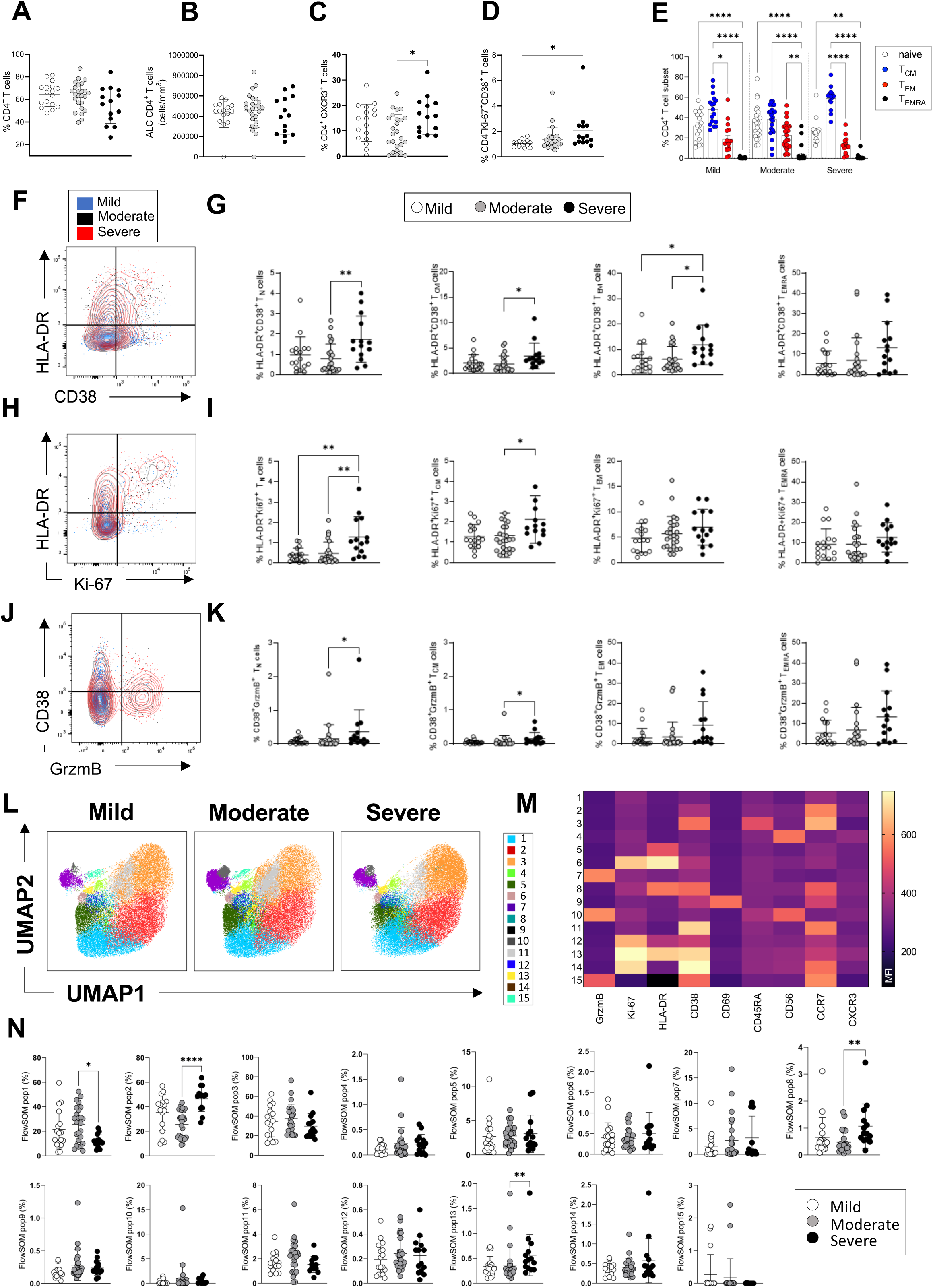
CD4^+^ T-cell profiles in convalescent COVID-19 patients at 3 months post admission. **A-D.** Percentage of CD4^+^ T-cells within the CD3^+^ gate (A), absolute number of CD4^+^ T-cells (cells/mm^3^) (B) and percentages of CD4^+^ T-cells expressing CXCR3 (C) and co-expressing Ki67/CD38 (D) are shown in mild, moderate and severe patients. **E.** Percentages of naïve (CCR7^+^ CD45RA^+^), T central memory (T_CM_, CCR7^+^ CD45RA^-^), T effector memory (T_EM_, CCR7^-^ CD45RA^-^), and T effector memory RA re-expressing (T_EMRA_, CCR7^-^ CD45RA^+^) CD4^+^ T-cells are shown for patients with mild, moderate and severe disease. **F.** Flow cytometry plot showing a representative staining from a mild, moderate and severe patient of HLA-DR and CD38 expression in CD4^+^ T_EM_ cells (overlaid and shown respectively in blue, black and red). **G.** Percentages of activated HLA-DR^+^CD38^+^ CD4^+^ T-cells within naïve, T_CM_, T_EM_ and T_EMRA_ cells. **H** Flow cytometry plot with a representative staining from a mild, moderate and severe patient of HLA-DR and Ki67 expression in CD4^+^ T_EM_ cells. **I.** Percentages of proliferating HLA-DR^+^ Ki67^+^ CD4^+^ T-cells within naïve, T_CM_, T_EM_ and T_EMRA_ cells. **J.** Flow cytometry plot with a representative staining from a mild, moderate and severe patient of HLA-DR and granzyme B (GrzmB) expression in CD4^+^ T_EM_ cells. **K.** Percentages of proliferating HLA-DR^+^ GrzmB^+^ CD4^+^ T-cells within naïve, T_CM_, T_EM_ and T_EMRA_ cells. **L.** Unsupervised UMAP analysis showing the FlowSOM clusters in mild (N=17), moderate (N=25) and severe (N=14) patients. Plots are gated on CD4^+^ T-cells. **M.** Heatmap with the expression of each analysed marker within the FlowSOM populations shown as Mean Fluorescent Intensity (MFI). **N**. Summary of the percentage of CD4^+^ T-cells within the indicated FlowSOM populations in mild, moderate and severe patients. Data in graphs are visualized as mean +/- SEM. Statistics are calculated by One-way ANOVA (Kruskal-Wallis test) with Dunn’s correction for multiple testing.

Similarly, the analyses of CD8^+^ T-cells showed comparable frequencies and absolute numbers of circulating CD8^+^ T-cells in patients across disease severities and increased levels of CD8^+^ T-cells expressing markers of activation (HLADR^+^CD38^+^) in severe compared to moderate patients and cytotoxicity (granzyme B^+^) in severe compared to mild patients (**Figure 2 A-D**). The overall differentiation status of CD8^+^ T-cells was comparable in patients across disease severities with detectable frequencies of naïve, T_CM_, T_EM_ and T_EMRA_ cells (**Figure 2 E**). CD8^+^ T-cells expressing markers of activation and cytotoxicity (HLADR^+^ CD38^+^ and CD38^+^ granzyme B^+^), could be detected at higher frequencies within T_CM_, T_EM_ and T_EMRA_ subsets in severe compared to moderate and/or mild patients (**Figure 2 F-I**). Unsupervised analysis by UMAP and FlowSOM of CD8^+^ T-cells showed the presence of 15 clusters of cells expressing the analyzed markers (**Figure 2 J, K**). The frequencies of a cluster of CD8^+^ T-cells expressing high levels of granzyme B and CD45RA (cluster 3) were significantly increased in both severe and moderate compared to mild patients and represented up to 50% of all CD8^+^ T-cells in severe/moderate groups and up to 30% in mild group (**Figure 2 L**). CD8^+^ T-cells expressing high levels of CCR7 and intermediate levels of granzyme B, CXCR3 and CD56 were increased significantly in both mild and severe compared to moderate patients (cluster 12). In summary, manual and unsupervised analysis revealed the presence of CD8^+^ T-cells expressing markers of activation and cytotoxicity which are increased in severe compared to moderate and/or mild patients. Analysis of expression of the same markers of activation/proliferation in NK CD56^dim^ and CD56^bright^ cells did not show evidence of ongoing NK cell activation and NK cells were similar in patients who recovered from mild, moderate and severe disease (**Supplementary Figure 3 A-F**). Similarly, analysis of the frequencies and expression of activation markers (CD80, CD68) of classical (CD14^+^ CD16^-^), intermediate (CD14^+^ CD16^+^) and non-classical (CD14^-^CD16^+^) monocytes showed similar frequencies of these cells across the disease severities and lack of ongoing activation (**Supplementary Figure 3 G-K**). Similar frequencies of unconventional TCR-γδ T-cells were detected in patients with mild, moderate and severe disease and these cells lacked expression of markers of activation/proliferation (**Supplementary Figure 3 L-N**). In contrast, CD19^+^ B cells from severe patients showed an increase in the expression of the activation marker CD80 compared to patients with mild disease, although expression of other markers of activation/proliferation did not differ (HLA-DR, CD38 and Ki67) (**Supplementary Figure 3 O-R**).

**Figure 2.**
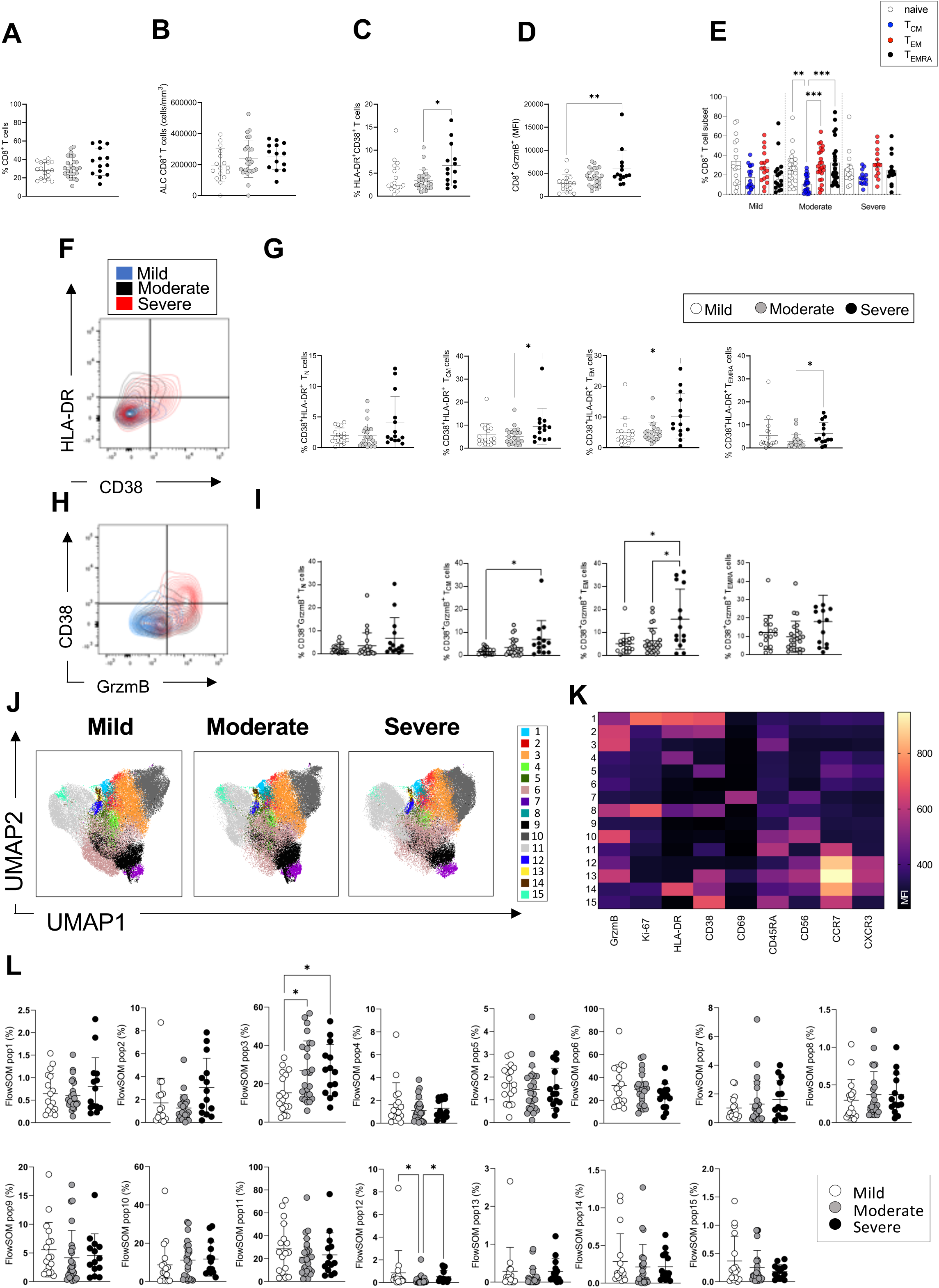
CD8^+^ T-cell profiles in convalescent COVID-19 patients at 3 months post admission. **A-D.** Percentage of CD8^+^ T-cells within the CD3^+^ gate (A), absolute number of CD8^+^ T-cells (cells/mm^3^) (B) and percentages of CD8^+^ T-cells co-expressing the activation markers HLA-DR/CD38 (C) or granzyme B (D, shown as MFI) are shown in mild, moderate and severe patients. **E.** Percentages of naïve (CCR7^+^ CD45RA^+^), T central memory (T_CM_, CCR7^+^ CD45RA^-^), T effector memory (T_EM_, CCR7^-^ CD45RA^-^), and T effector memory RA re-expressing (T_EMRA_, CCR7^-^ CD45RA^+^) CD8^+^ T-cells in patients with mild, moderate and severe disease. **F.** Flow cytometry plot with a representative staining from a mild, moderate and severe patient (overlaid and shown respectively in blue, black and red) of HLA-DR and CD38 expression in CD8^+^ T_EM_ cells. **G.** Percentages of activated HLA-DR^+^ CD38^+^ CD8^+^ T-cells within naïve, T_CM_, T_EM_ and T_EMRA_ cells. **H.** Flow cytometry plot with a representative staining from a mild, moderate and severe patient of HLA-DR and granzyme B (GrzmB) expression in CD8^+^ T_EM_ cells. **I.** Percentages of proliferating HLA-DR^+^ GrzmB^+^ CD8^+^ T-cells within naïve, T_CM_, T_EM_ and T_EMRA_ cells. **J.** Unsupervised UMAP analysis showing the FlowSOM clusters in mild (N=17), moderate (N=25) and severe (N=14) patients. Plots are gated on CD8^+^ T-cells. **K.** Heatmap with MFI levels for each analysed marker within the FlowSOM populations. **L**. Summary of percentage of CD8^+^ T-cells within the indicated FlowSOM populations in mild, moderate and severe patients. Data in the graphs are shown as mean +/- SEM. Statistics were calculated by One-way ANOVA (Kruskal-Wallis test) with Dunn’s correction for multiple testing.

In summary, both manual and unsupervised analysis showed increased frequencies of activated/proliferating CD4^+^ and CD8^+^ T-cells in severe compared to mild and/or moderate patients at 3 months post admission, suggesting the presence of on-going immune activation in these patients. We did not find significant activation of other immune cells analysed (NK, TCR-γδ, monocytes), with the exception of CD19^+^ B cells from severe patients which expressed increased levels of CD80 compared to mild patients.

### Elevated pro-inflammatory cytokines/chemokines in severe patients at 3 months

To gain insights into the factors that may be driving CD4+ and CD8+ T-cell activation and proliferation and define the nature of any persistent inflammation in these patients, we investigated the presence of soluble circulating pro-inflammatory cytokines/chemokines in the plasma of COVID-19 patients 3 months after admission. The plasma levels of the following 23 cytokines/chemokines were measured using a Luminex platform: GM-CSF, IFN-γ, IFN-α, IL-1α, IL-1β, IL-10, IL-12 p70, IL-13, IL-15, IL-17A, IL-18, IL-2, IL-4, IL-5, IL-6, IL-7, IL-8, IP-10, MCP-1, MIP-1α, MIP-1β and TNF-α (Human Procarta plex). Our results show that at 3 months the levels of IL-4, IL-7, IL-17 and TNF-α were significantly increased in the plasma of patients with severe compared to mild and/or moderate disease, while IFN-γ was increased in moderate compared to mild patients. Interestingly, IFN-γ was largely undetectable in the plasma of severe patients (**Figure 3 A**). A trend for increased levels of IL-18 was observed in the plasma of severe compared to mild and moderate patients but the difference was not significantly different. The plasma levels of other cytokines/chemokines tested did not differ between patients with different disease severities at 3 months (**Supplementary Figure 4**). We next investigated the kinetics of expression of these cytokines in patients at 3, 8 and 12 months post admission and observed a statistically significant decrease of plasma levels of IL-4, IL-12, IL-13, and TNF-α from 3 to 8 and/or 12 months, suggesting that at 3 months the levels of these cytokines may still be affected by events occurring during the acute phase of infection. IL-15 levels were similar at 3 and 8 months and decreased thereafter (**Figure 3 B, C**). IL-17 levels were higher at 3 months and progressively diminished over time, although differences were not statistically significant. In contrast, IP-10 levels were higher at 8 and 12 months compared to 3 months. The plasma levels of the other cytokines analysed including IL-7, IFN-γ, IL-18 and MCP-1 were comparable at 3-12 months post admission. Based on our results in Figures 1-2 showing increased levels of circulating activated/proliferating CD4^+^ and CD8^+^ T-cells in severe patients and on the elevated levels of cytokines involved in T-cell proliferation (IL-7, IL-15, TNF-α) at 3 months, we asked whether factors present in the plasma of severe patients might render T-cells more susceptible to “bystander T-cell” activation, which occurs during viral infection and is believed to be driven by IL-15. To address this, we co-cultured purified T-cells from healthy donors (n=4) for 7 days with plasma derived from either heterologous healthy donors (n=4, “HD plasma”) or patients with mild or severe COVID-19 (n=4, “Mild plasma”; n=4, “Severe plasma”). We observed a significant upregulation of the IL-15Rα−chain, the IL-15R subunit that binds directly to IL-15, in T-cells co-cultured in the presence of plasma from severe patients compared to healthy donors (**Figure 3D, E**). These experiments suggest that 3 months after severe COVID-19, peripheral blood CD4^+^ and CD8^+^ T-cells are exposed *in vivo* to plasma factors that make them more responsive to IL-15 by inducing upregulation of the IL-15R, and that persistent T-cell activation observed *ex vivo* at 3 months may be driven by cytokines such as IL-15.

**Figure 3.**
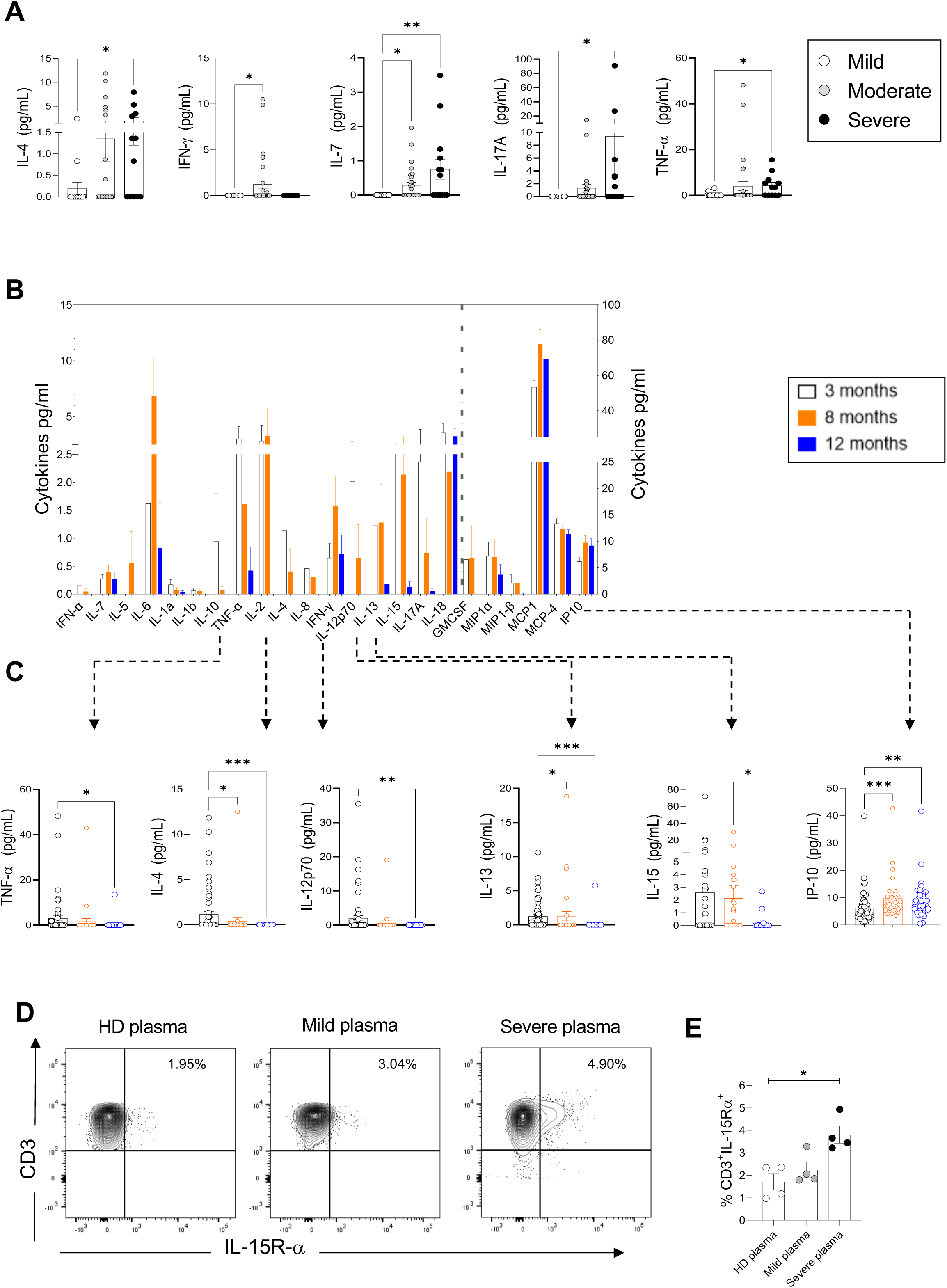
Plasma pro-inflammatory cytokines/chemokines measured at 3, 8 and 12 months. **A.** Plasma cytokines/chemokines measured at 3 months post admission which differed significantly between patients with mild, moderate and severe disease are shown (N=63: Mild: N=17; Moderate: N=32; Severe: N=14, depicted in white, grey and black bars, respectively). **B-C.** Cytokines/chemokines measured longitudinally in matched samples in patients at 3 (n=63), 8 and 12-months post admission (n=33 samples for each time point) are shown. Data from analytes that differed significantly between time points in B are shown in C for each patient. **D, E**. Purified CD3^+^ T-cells from 4 healthy donors were co-cultured with plasma from 4 healthy donors, 4 mild and 4 severe patients. Shown is IL-15R-α expression in T-cells from a representative donor (D) and the average expression of IL-15Rα by T-cells from each PBMC donor after co-culture with plasma from healthy, mild and severe patients, where each data point represents a single patient (E). Statistics were calculated by One-Way ANOVA test (Kruskal-Wallis test) with Dunn’s multiple comparison test (A, E) and by Anova/repeated-measures one-way Anova, mixed-effects analysis with the Geisser-Greenhouse correction, Tukey’s multiple comparison test. (B, C). Data are visualized as mean with +/- SEM. Measurements for each patient/donor in A-C and E are performed in technical duplicates.

In summary, at 3 months post admission we observe increased levels of IL-4, IL-7, IL-17 and TNF-α in the plasma of severe compared to mild and/or moderate patients. The levels of IL-4, TNF-α and IL-17 gradually diminished from 3 to 12 months suggesting a resolution in the perturbations of immune cells that are secreting these cytokines, while those of IL-7 where comparable across these timepoints.

### Robust SARS-CoV-2 memory T-cell and antibody responses

To investigate the magnitude and functional features of SARS-CoV-2-specific T-cells and address potential associations with disease severity and long COVID, we performed IFN-γ Enzyme-Linked Immune absorbent Spot (ELISpot) assays with PBMC samples from 61 patients. IFN-γ production by ELISpot was measured in PBMCs after an overnight stimulation with overlapping 15-mer peptide pools spanning the sequences of the SARS-CoV-2 structural proteins spike (divided into 2 pools: spike 1 and 2, which include peptides spanning the N and C terminal regions of spike, respectively), membrane and nucleocapsid. To investigate in parallel the T-cell response to a persistent virus that is commonly present in the UK population, PBMCs were stimulated with a 15-mer peptide pool spanning the human cytomegalovirus (CMV) pp65 protein. Positive (PMA/ionomycin) and negative control wells were included in the assay. The ELISpot results showed that the memory T-cell response to SARS-COV-2 spike, membrane and nucleocapsid was robust and comparable in mild, moderate and severe patients at 3 months post admission. T-cell responses targeting CMV and polyclonal T-cell activation induced by PMA/ionomycin were also similar in mild, moderate and severe patients (**Figure 4 A-F**). The percentage of “responders” to SARS-CoV-2, calculated as the percentage of patients who presented a response of >5 SFC/10^5^ PBMCs to peptides spanning the indicated protein sequence, was higher in moderate versus severe patients but similar between mild and moderate patients (**Figure 4 G**). We also assessed the capacity of mild, moderate and severe patients to mount an antibody response targeting SARS-CoV-2 by using a highly sensitive plate-based luciferase immunoprecipitation assay to the spike receptor binding domain (RBD), which detects all antibody isotypes. SARS-CoV-2 antibody responses were detectable across all patient groups but were higher in patients with moderate compared to mild disease (**Figure 4 H**).

**Figure 4.**
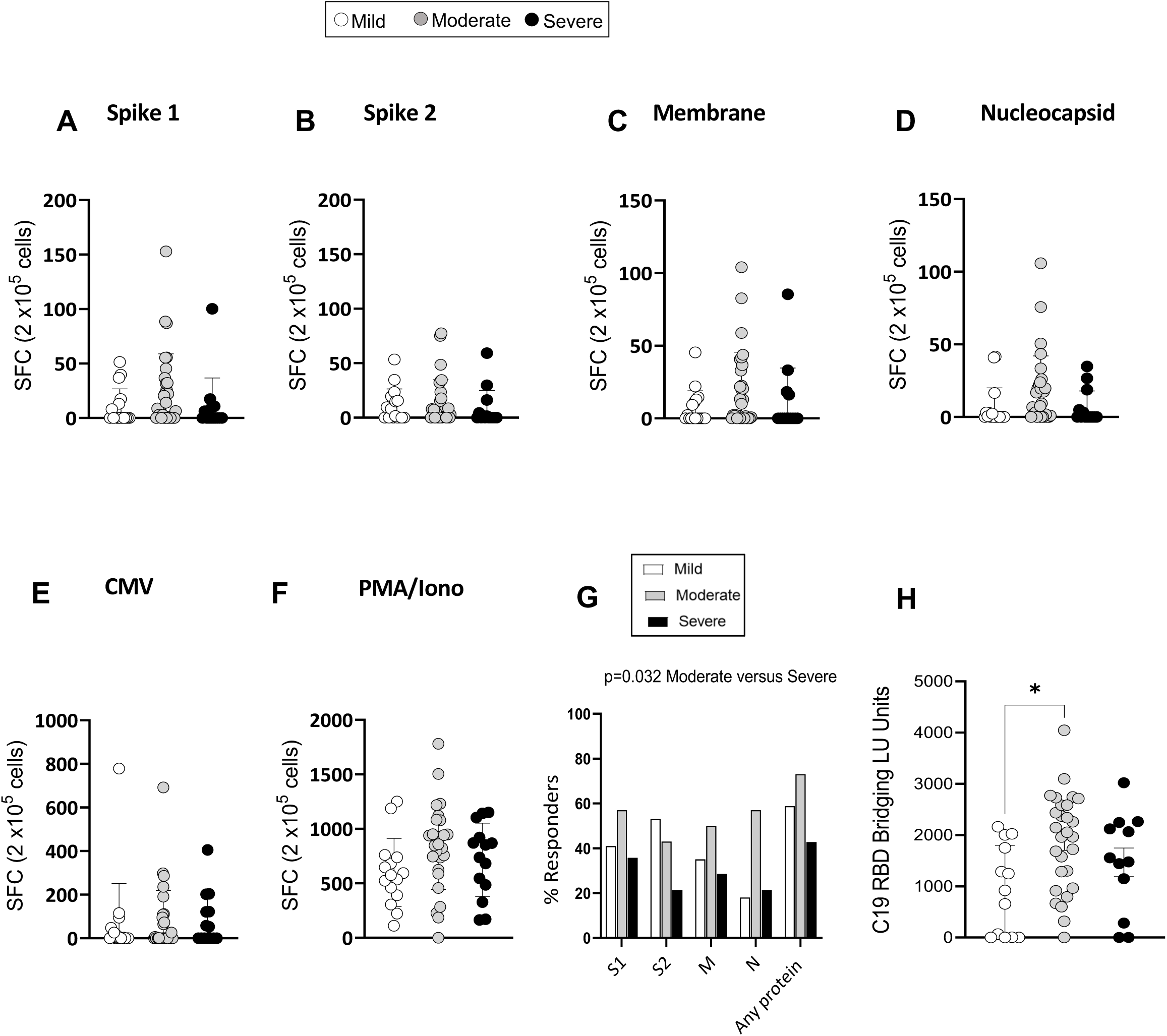
SARS-CoV-2-specific memory T-cell and antibody response at 3 months. **A-F.** IFN-γ release measured by ELISpot in PBMCs from mild, moderate and severe patients (N=61) upon stimulation with 15-mer peptide pools spanning SARS-CoV-2 spike 1 (A), spike 2 (B), membrane (C), nucleocapsid (D), CMV pp65 (E) and PMA/ionomycin (F). Results are shown as Spot Forming cells (SFC) relative to 2x10^5^ PBMCs. **G**. Percentages of responders assessed as patients from each severity group who displayed a response to the indicated peptide pool >5 SFC/2x10^5^ PBMCs**. H.** SARS-CoV-2 RBD antibody titers in patients expressed as RBD Bridging LU units. Data in A-F are visualized as mean +/- SEM. Measurements for each patient are performed in technical duplicates. Statistics were calculated by One-way ANOVA (Kruskal-Wallis test) with Dunn’s correction for multiple testing.

In summary, the overall magnitude of IFN-γ producing memory T-cells targeting SARS-CoV-2 spike, membrane and nucleocapsid assessed by ELISpot was comparable between mild, moderate and severe patients at 3 months post admission. However, the proportion of patients who displayed a T-cell response to SARS-CoV-2 peptides (responders) was higher in moderate compared to severe patients. The titers of RBD-specific antibodies were also higher in moderate compared to mild patients, suggesting a superior capacity of patients with moderate disease to mount detectable memory T-cell and antibody responses to SARS-CoV-2.

### Lower frequencies of IFN-γ^+^/TNF-α^+^ and/or CD107a^+^ SARS-CoV-2-specific T-cells in mild patients

To evaluate the relative contribution of CD4^+^ and CD8^+^ T-cells to the SARS-CoV-2 T-cell response observed by ELISpot and to evaluate the cytokine profiles and phenotype of SARS-CoV-2-specific CD4^+^ and CD8^+^ T-cells in these patients in-depth, we performed intracellular cytokine staining (ICS) by flow cytometry in samples from 39 patients for which we had available PBMC samples, at 3 months post admission. PBMCs were briefly stimulated with or without SARS-CoV-2 peptides spanning the spike protein or with PMA/ionomycin as a positive control, and cells were subsequently stained with antibodies recognizing T-cell markers of differentiation (CD45RA, CCR7), activation (HLA-DR, CD38), proliferation (Ki67) and tissue-homing (CXCR3) and assessed for production of IFN-γ, TNF-α and IL-2. Representative flow cytometry plots showing production of IFN-γ, TNF-α and CD107a are shown in **Figure 5 A**. Due to limited cell numbers we were able to comprehensively assess in all samples the T-cell response to only spike-1 peptide pool. Results following spike-2 peptide stimulations were similar to those obtained with spike-1, but were available for fewer patients (data not shown). Our results show that frequencies of spike-1-specific CD4^+^ memory T-cells producing IFN-γ and/or TNF-α or IFN-γ and/or CD107a upon encounter of SARS-CoV-2 spike-1 peptides were consistently higher in moderate compared to mild patients. In contrast CD8^+^ T-cells producing IFN-γ and/or TNF-α were similar across the disease severities and those producing IFN-γ and/or CD107a were higher in moderate and severe compared to mild patients (**Figure 5 B**). The polyfunctionality of T-cells, defined as their ability to simultaneously perform more than one function, has been associated with better viral control in chronic viral infections such as HIV. We therefore investigated the polyfunctionality of CD4^+^ and CD8^+^ T-cells specific for SARS-CoV-2 spike peptide pools. Across all patient groups monofunctional T-cells performing a single function dominated the response, representing 66-91% of total spike-1-specific T-cells, followed by T-cells performing 2 or 3 functions. T-cells performing 4 functions (IFN-γ^+^ IL-2^+^ TNF-α^+^ CD107a^+^) were largely absent (**Figure 5C, D,** and data not shown for IL-2). Across all patient groups monofunctional CD4^+^ and CD8^+^ T-cells targeting spike-1 peptide pools were significantly higher than their polyfunctional counterparts (**Figure 5 E**), suggesting that T-cell functionality of SARS-CoV-2 memory T-cells is not influenced by disease severity. Spike-1-specific CD4^+^ T-cells were mainly contained within the T_EM_ population, while Spike-1-specific CD8^+^ T-cells were contained within T_EM_ and T_EMRA_ populations (**Figure 5 F**), and results were similar between patient groups. CXCR3 expression was also similar in CD4^+^ and CD8^+^ T-cells across patient groups (data not shown).

**Figure 5.**
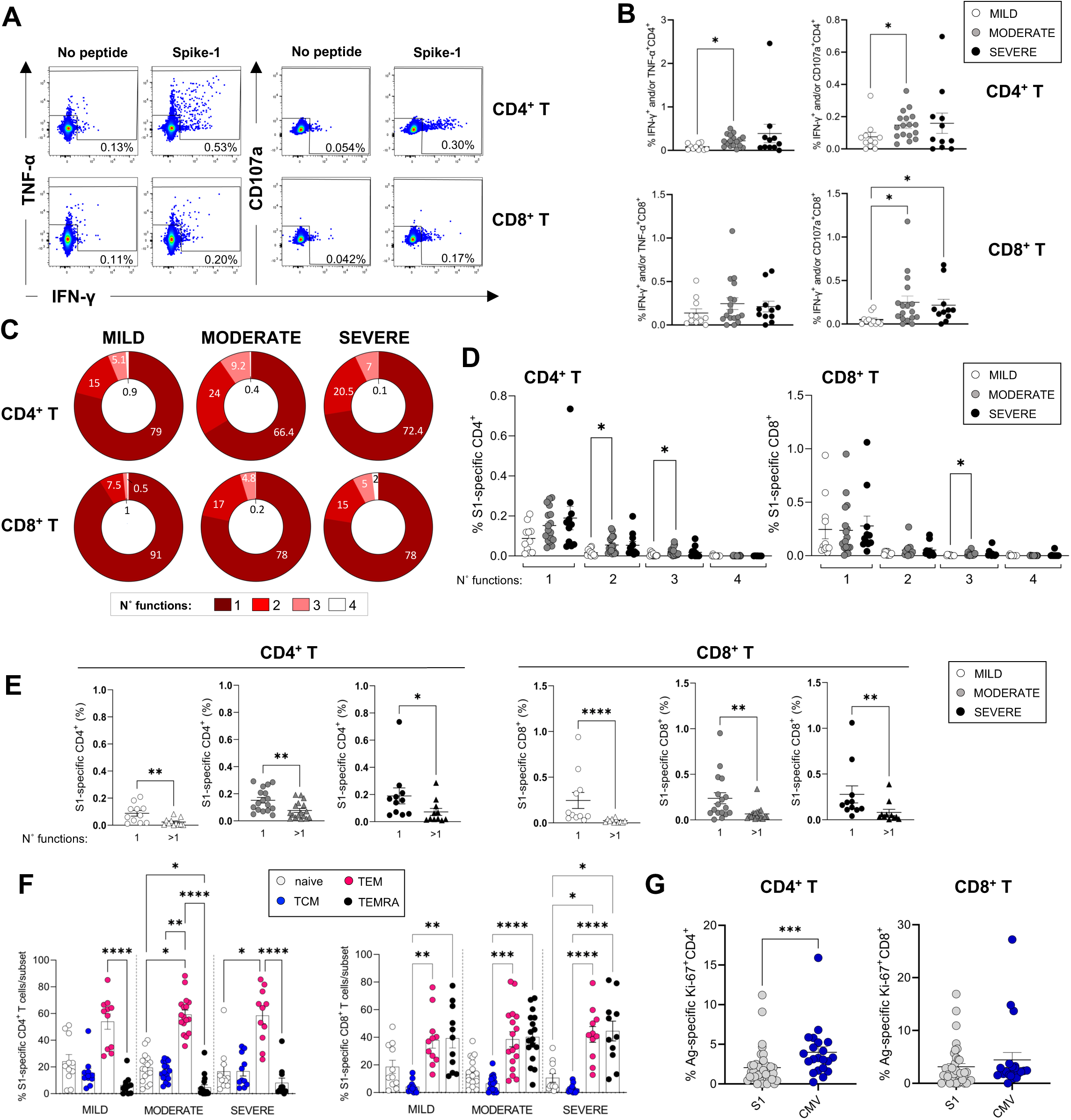
Magnitude and cytokine profiles of SARS-CoV-2-specific CD4^+^ and CD8^+^ T-cells at 3 months. CD4^+^ and CD8^+^ T-cell responses targeting spike peptides were assessed by ICS in mild, moderate and severe patients. **A, B.** Shown are representative flow cytometry plots of IFN-γ and TNF-α or CD107a production by CD4^+^ and CD8^+^ T-cells (A) and the percentages of CD4^+^ (top panel) and CD8^+^ T (bottom panel) cells producing IFN-γ and/or TNF-α and IFN-γ and/or CD107a in the presence of spike-1 peptides (B). **C.** Pie charts summarizing the multifunctionality of T-cells specific for spike-1, defined as their capacity to produce 1, 2, 3 or 4 cytokines/CD107a (n° functions). **D.** Spike-1 (S1) specific CD4^+^ (left panel) and CD8^+^ T-cells (right panel) that express 1-4 functions in mild, moderate and severe patients. **E.** Monofunctionality and polyfunctionality (>1 function) of CD4^+^ and CD8^+^ T-cells targeting spike-1 peptides in mild, moderate and severe patients. **F.** Expression of differentiation markers CD45RA/CCR7 by spike-1 specific CD4^+^ and CD8^+^ T-cells in mild, moderate and severe patients. Naïve cells= CCR7^+^CD45RA^+^ (white); T central memory cells (TCM)= CCR7^+^ CD45RA^-^ (blue); T effector memory cells (T_EM_)= CCR7^+^ CD45RA^-^ (red); T effector memory RA re-expressing cells (T_EMRA_)= CCR7^+^ CD45RA^-^ (black). **G.** Percentage of Spike-1-specific or CMV-specific CD4^+^ (left panel) and CD8^+^ T (right panel) cells that express Ki67. Data in A-B, D-G are visualized as mean +/- SEM. Statistics were calculated by One-way ANOVA (Kruskal-Wallis test) with Dunn’s correction for multiple testing or by Mann Whitney t-test.

We next asked whether we could detect any ongoing activation and/or proliferation of SARS-CoV-2 or CMV-specific T-cells at 3 months post admission to understand whether these cells contributed to the total pool of activated/proliferating cells that were increased in severe patients. Our results showed that CMV-specific CD4^+^ but not CD8^+^ T-cells expressed Ki67 at significantly higher levels compared to their Spike-1-specific counterparts, suggesting that CMV-specific CD4^+^ T-cells were proliferating albeit at low levels (**Figure 5 G**). These data suggest that CMV-specific CD4^+^ T-cells may contribute to the pool of activated CD4^+^ T-cells detected in peripheral blood of COVID-19 patients at 3 months.

In summary, SARS-CoV-2-specific CD4^+^ and CD8^+^ T-cells could be detected in the peripheral blood of patients 3 months after mild, moderate and severe COVID-19. An in-depth analysis of SARS-CoV-2-specific T-cells producing IFN-γ, TNF-α, IL-2 and CD107a revealed an increased frequency of T-cells targeting spike-1 in moderate and/or severe compared to mild patients.

### Associations between on-going symptoms and immune response

At the 3 month follow-up clinic, patients were screened for a pre-defined list of symptoms in a clinician-led clinic, as reported elsewhere.^33^ Of all patients included in this study, 79% (82%, 75% and 86% of mild, moderate and severe patients, respectively) reported at least one ongoing symptom with breathlessness and excessive fatigue being the most common. Symptoms also included muscle weakness, sleeping difficulty, psychiatric symptoms, anosmia, chest pain and cough (**Table 1**). Patients who recovered from severe disease experienced each of these symptoms more frequently compared to patients from the mild and moderate groups, except for cough and chest pain which were reported by a minority of patients across all groups. The proportion of patients who did not report any symptoms was largely similar in mild, moderate and severe patients (**Figure 6 A**). Mild patients more frequently reported 1 symptom, while those from the severe group were more likely to report 4 symptoms. Moderate patients reported 0-2 symptoms with similar frequencies (**Figure 6 B**).

**Figure 6.**
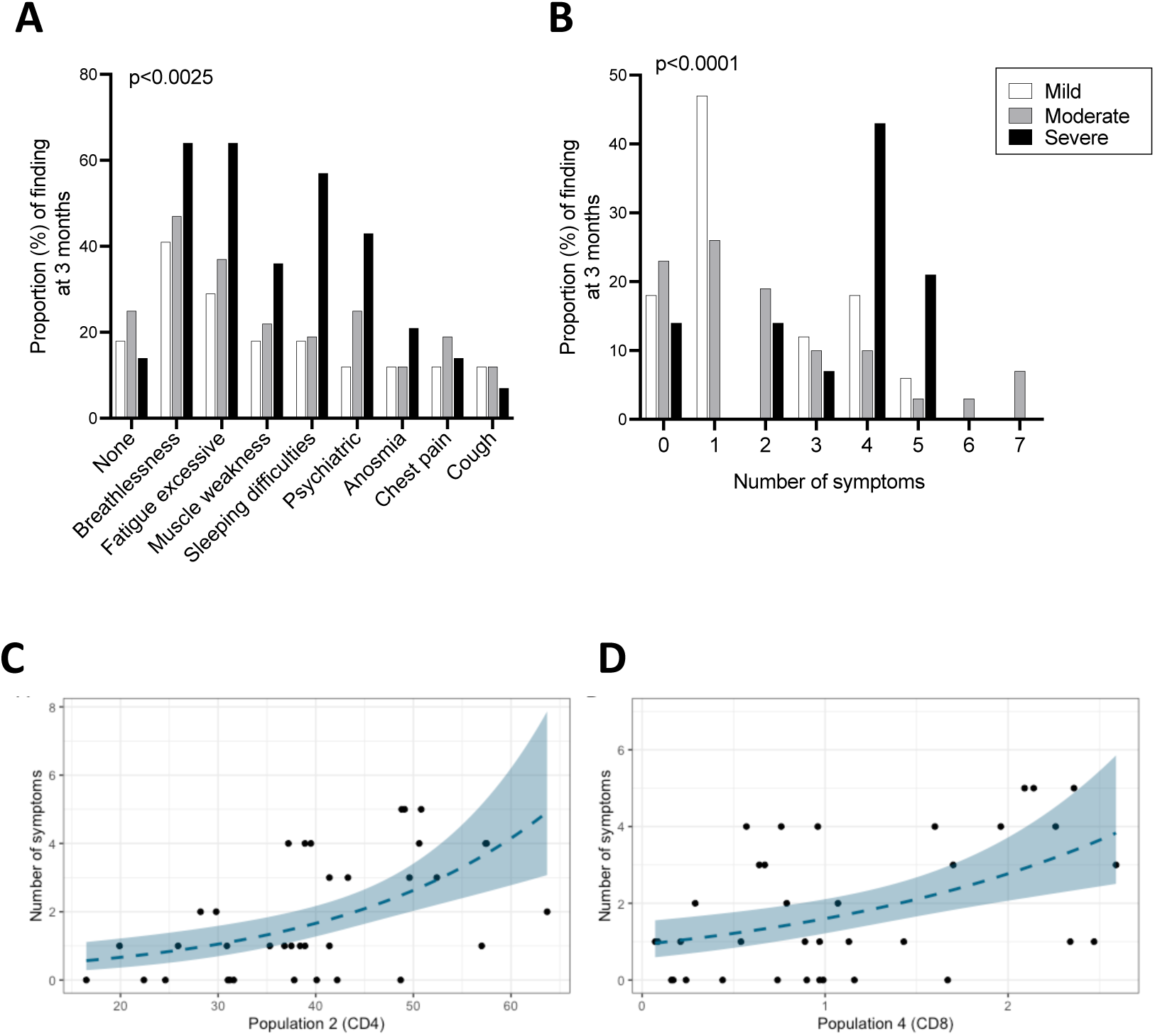
Ongoing symptoms at 3 months and associations with immune profiles. **A,B.** The percentage of mild, moderate and severe COVID-19 patients who reported the indicated symptom (A) or number of symptoms (B) at the 3 months are indicated with white, grey and black bars, respectively. Statistics were calculated using a Chi-square test. **C, D.** Graphs depicting the association between number of symptoms and UMAP T-cells clusters in Poisson models, specifically CD4^+^ T-cell cluster 2 (C) and CD8^+^ T-cell cluster 4(D).

We next asked whether ongoing symptoms at 3 months associates with T-cell profiles and/or the plasma pro-inflammatory cytokines assessed in this study. Poisson regression was performed for the total number of symptoms reported at 3 months against a set of immune parameters of interest including cytokines/chemokines that could be detected consistently across patients (IFN-γ, IL-12p70, IL-13, IL-15, IL-17A, IL-18, IL-4, IL-7, IP-10 and TNF-α) and T-cell profiles detected by flow cytometry (populations identified by manual gating and UMAP/FlowSOM). In the analyses that were unadjusted for sex, age and severity the number of long COVID symptoms were found to associate significantly with two clusters of T-cells that were identified in the FlowSOM analyses (CD4^+^ T-cell cluster 2: FDR p=0.003; CD8^+^ T-cell cluster 4: FDR p=0.015). CD4^+^ T-cell cluster 2 represents a population of cells present at higher frequencies in severe compared to moderate patients, which expressed CCR7, CD38 and intermediate/low levels of Ki67, suggesting moderate/recent activation (**Figure 2 N**). These cells represent on average approximately 50% of all CD4^+^ T-cells in severe patients. In contrast, CD8+ T-cell cluster 4 is present in similar frequencies across disease severities and represents a subset of cells that express HLA-DR, intermediate levels of CCR7 and CXCR3 and lacks granzyme B expression (**Figure 3 K**). Other immune parameters including the frequencies of activated/proliferating CD4^+^ T-cell subsets and UMAP CD4^+^ T-cell cluster 11 (CD38 high, Ki67 intermediate CD4^+^ T-cells) associated inversely with the number of symptoms with uncorrected p-values <0.05, but these associations were not statistically significant after FDR correction. The ratio of CD4^+^/CD8^+^ T-cells correlated directly with the number of symptoms but similarly the association was not significant after FDR correction (**Supplementary Table 2A**). In an analysis adjusted for sex, age and severity none of the estimates displayed FDR p<0.05. FlowSOM CD4^+^ T-cell cluster 2 and CD8^+^ T-cell cluster 4 that significantly associated with the number of symptoms in the unadjusted analysis failed to associate significantly after adjusting for age, sex and severity. Other immune parameters including plasma cytokine levels of IP10, IL-4, IFN-γ and IL-12, UMAP CD8^+^ T-cell cluster 8 and frequencies of Ki67^+^ HLA-DR^+^ CD4^+^ T_EM_ cells directly correlated with number of symptoms (p<0.05), but p-values were >0.05 after FDR correction (**Supplementary Table 2 A**). We also did not identify any association with T-cell profiles or cytokines/chemokines and the physical and mental composite scores.

## Discussion

Emerging data demonstrates that immune activation may persist for months after COVID-19, however there is heterogeneity in findings particularly between hospitalized versus non-hospitalized cohorts, which by nature of how patients were managed during the peak of the pandemic comprise of patients with a very different spectrum of disease. Here we report results from a small study performed on a cohort of clinically well-characterized mild, moderate and severe patients who were all hospitalized at the start of the pandemic and followed up closely through outpatient follow-up clinics up to 12 months post admission. We ask whether ongoing inflammation and immune perturbations, particularly within the T-cell population, associate with long COVID symptoms. Specifically, we ask the following questions: (1) Is there evidence of ongoing inflammatory events post hospitalization with COVID-19, and are these linked with disease severity at acute infection? (2) Are the magnitude and features of the SARS-CoV-2-specific CD4^+^ and CD8^+^ T-cell memory response influenced by disease severity? (3) Do long COVID symptoms associate with persistent immune activation, inflammation and/or SARS-CoV-2-specific T-cell immunity?

To answer these questions, we performed an in-depth immunological analysis of 154 samples from 63 patients at 3 months post admission, including a characterization of activation/proliferation profiles and phenotypic features of circulating CD4^+^ and CD8^+^ T-cells, γδ-T-cells, NK cells, B cells and CD14^+/-^CD16^+/-^ monocytes by flow cytometry, as well as an analysis of T-cells and antibodies targeting SARS-CoV-2 and pro-inflammatory plasma cytokines/chemokines by Luminex at 3, 8 and 12 months. Our data shows persistent CD4^+^ and CD8^+^ T-cell activation and elevated plasma levels of IL-4, IL-7, IL-17 and TNF-α at 3 months, in severe compared to mild and/or moderate patients. The elevated levels of these cytokines are consistent with ongoing T-cell activation as CD4^+^ T-cells are key producers of IL-4 and IL-17 and both CD4^+^ and CD8^+^ T-cells secrete TNF-α. IL-7 stimulates proliferation of T-cells and can be produced by a variety of cells such as epithelial cells, stromal cells and dendritic cells. Plasma levels of IL-4, IL-7 and IL-15 were similar at acute infection and 3 months but declined at later time points, suggesting a slow recovery of T-cell related perturbations within 12 months. CD4^+^ and CD8^+^ T-cell activation was evident when we analyzed the flow cytometry data both by manual gating and by FlowSOM/UMAP - where the former method investigates cells expressing known combinations of markers, while the latter identifies clusters of cells in an unsupervised manner based on the combined expression of all markers analyzed. Manual gating highlighted increased levels of CD4^+^ T-cells expressing the lung tissue-homing marker CXCR3 in severe patients, suggesting the presence of factors that may be recruiting and priming CD4^+^ T-cells in this site. This finding may suggest unresolved damage/inflammation in the lung, however clinical abnormalities in lung function were evident only in a minority of these patients during the follow-up clinic.^33^ Both manual and FlowSOM/UMAP analysis show increased granzyme B expression in CD8^+^ T-cells from severe patients, suggesting an overall higher cytotoxic ability of these cells. Granzyme B expression is upregulated in cells after T-cell receptor (TCR) stimulation and confers higher cytotoxic abilities to the activated T-cell, hence higher expression could reflect a higher activation state in these cells. Ongoing T-cell activation did not appear to impair the ability of severe patients to mount and maintain memory T-cell or antibody responses to SARS-CoV-2 structural proteins, which were present and robust across all patients. The quality of the T-cell response was also similar across the patient groups, and we could detect monofunctional and polyfunctional SARS-CoV-2-specific T-cells, with the former dominating the response. However, moderate compared to mild patients displayed elevated spike RBD antibody titers and higher magnitudes of CD4^+^ and CD8^+^ T-cells producing IFN-γ and/or TNF-α/CD107a by ICS, after stimulation with spike-1 peptides. The magnitude of the T-cell response is closely linked with viremia^34^ and higher viral loads during the acute illness may underlie the higher magnitude of IFN-γ/TNF-α producing CD4^+^ and CD8^+^ T-cells. However, viremia data at acute illness was not available for our patient cohort. The frequency of T-cells producing IFN-γ alone did not differ in the ICS analysis, in line with the ELISpot data. Cytokine production following PMA/ionomycin were comparable between patients, suggesting similar T-cell functionality to polyclonal stimulation that bypasses the TCR.

T-cells can be activated through TCR triggering or “bystander activated” by cytokines such as IL-7 and IL-15. Bystander T-cell activation was observed to occur during viral infection and is likely driven by IL-15.^35, 36^ Here we demonstrate that co-culture of plasma from severe patients induced upregulation of the IL-15 receptor (IL-15R)-α chain on T-cells from healthy donors, suggesting that the low-level proliferation of T-cells we see in severe patients may be bystander in nature and driven by plasma cytokines persistently present at 3 months in severe patients. Highly differentiated T-cells are more responsive to IL-15 due to their higher expression of the IL-15R.^37, 38^ The higher responsiveness of CMV-specific T-cells, which are highly differentiated cells has been proposed as a mechanism to explain why these cells are commonly activated/proliferating during acute viral infections such as dengue, HBV and COVID-19.^35, 36, 39^ It may be possible that the low-level proliferation of CMV-pp65-specific CD4^+^ T-cells observed in this study is driven by cytokines which are still elevated at 3 months. Our data does not support the persistence of SARS-CoV-2 antigens at 3 months for two reasons. Firstly, we would expect that the persistence of SARS-CoV-2 antigens *in vivo* would result in the activation of SARS-CoV-2-specific T-cells, however we did not observe expression of markers of activation nor proliferation in spike-1-specific T-cells in our ICS analysis. Secondly, we did not detect any SARS-CoV-2 RNA by RT-PCR – however we cannot exclude that SARS-CoV-2 antigens may still be present but are sequestered in specific sites to which T-cells do not have access.

In this study the levels of CRP, albumin and IL-6 at 3 months were similar across patients, suggesting an overall resolution of the inflammatory processes occurring during the acute infection. In contrast, a recent study showed increased levels of several pro-inflammatory markers at 12 months, including CRP and IL-6 which correlated with the presence of persistently activated T-cells. Interestingly the authors showed that COVID-19 mRNA vaccination and boosting of SARS-CoV-2 immunity did not alter these profiles, suggesting inflammation may be independent of the presence of SARS-CoV-2 antigens in these patients.^17^ We cannot exclude that the presence of activated T-cells may be linked to the comorbidities experienced by the patients in this cohort, which included obesity, type-1 and 2 diabetes and asthma. These co-morbidities were present across all groups but increased progressively with disease severity (Table I). Long COVID symptoms were reported in 80% of patients across all disease severities but were more frequent in patients with severe disease, in line with published findings.^40^ The factors driving long COVID remain largely unclear, as do those underlying the long-term fatigue syndromes that have previously been observed following other viral infections.^41^ Although the prevalence of long COVID is reported to be higher in hospitalized patients and those who received intensive care support, the risk factors of developing a severe acute illness (male sex, age, obesity, ethnicity and cardiovascular disease) only partially overlap with those reported to associate with long COVID (female sex, age, obesity, anxiety, asthma), suggesting that pathogenesis at acute infection and in convalescence may be driven by distinct mechanisms.^25,26,27,28^ In this study, Poisson regression analysis showed no association between ongoing symptoms and immune perturbations after adjusting for age, sex and severity grades, but identified a significant association between ongoing symptoms and the frequency of a cluster of moderately activated CD4^+^ T-cells (cluster 2) and a cluster of activated, non-cytotoxic HLA-DR^+^ CD8^+^ T-cells (cluster 4) in unadjusted analyses, after FDR correction. Our study adds to emerging data suggesting that a prolonged immune activation can be observed following COVID-19 which may not directly associate with long COVID.^25, 26^ The clinical implications of persistent T-cell activation following COVID-19 remain unclear and warrant further investigation.

In summary, our study highlights a complex recovery of the immune sysT_EM_ following severe COVID-19 with evidence of persistent activation of CD4^+^ and CD8^+^ T-cells, which may be bystander driven, and elevated levels of T-cell-related cytokines in the plasma of severe patients at 3 months. Our data suggests the lack of a direct association between long COVID and immune activation markers and pro-inflammatory cytokines measured in this study.

## Data Availability

Flow cytometry data will be deposited in FlowRepository upon acceptance of this manuscript for publication; codes used for analysis (tidyverse package) will be provided upon request

## Acknowledgments

The authors wish to acknowledge the assistance of Dr Andrew Herman, Helen Rice, Poppy Miller and the University of Bristol Faculty of Biomedical Sciences Flow Cytometry Facility as well as Dr Kapil Gupta at the School of Biochemistry for Spike-RBD protein and members of the Diabetes and Metabolism team for help in testing samples for Spike-RBD antibodies. We would also like to thank the Bristol UNCOVER team for helpful discussions during the execution of this work and preparation of the manuscript. We are grateful to all patients and their families for participating in this study. This work was supported by the Elizabeth Blackwell Institute (EBI) with funding from the University’s alumni and friends (TRACK award to L.R. and grants to L.W and A.G) and by Southmead Hospital Charity (Registered Charity Number: 1055900).

## Material and Methods

### Patients

Patients hospitalized with COVID-19 (≥18 years of age) were recruited between 30^th^ March and 3^rd^ June 2020 into the observational study DIagnostic and Severity markers of COVID-19 to Enable Rapid triage (DISCOVER), a single-centre prospective study based in Bristol (UK). Ethics approval: REC:20/YH/1021. Survivors were invited at 3, 8 and 12 months post admission to attend outpatient follow up clinics for a systematic clinical assessment (Arnold et al 2020). For those patients attending a face-to-face follow-up, consent was taken to collect samples for research purposes (blood for PBMC isolation, plasma and serum). When available serum collected from patients at admission was made available to the research team.

### RT-PCR analysis

RT-PCR analysis was performed on all plasma samples used for Luminex for biosafety reasons (acute, 3-12 months). SARS-CoV-2 test was performed by an in-house RT-PCR at the regional Southwest Public Health England Regional Virology laboratory, utilising a PHE approved assay at the time of testing. All plasma samples were RT-PCR negative except for 2 samples taken at acute infection.

### Isolation of peripheral blood mononuclear cells (PBMCs)

Blood samples were collected from COVID-19 patients after informed consent in EDTA vacutainer tubes and Peripheral blood mononuclear cells (PBMCs) and plasma were isolated from peripheral blood using Leucosep tubes containing Ficoll and cryopreserved. PBMCs were resuspended in freezing media (10% DMSO, 90% FCS) prior to freezing in -80°C and frozen vials were transferred to liquid nitrogen 24-48 hours later.

### T-cell co-culture with patient plasma

PBMCs were isolated from the blood of healthy donors (N=4) and CD3^+^ T-cells were isolated with magnetic beads using Pan T-cell Isolation Kit, (Miltenyi), according to the manufacturer’s instructions. Purified CD3^+^ T-cells from healthy donors were then labelled with CellTrace™ Violet (Thermo Fisher) and 3 x10^5^ purified T-cells were co-cultured in round-bottom 96-well plate with plasma derived from either a heterologous healthy donor (N=4 healthy plasma), severe (N=4 severe plasma) or mild patients (N=4 mild plasma) for 7 days. Each condition was performed in technical duplicates. Cells were also plated in the presence of anti CD3/CD28 Dynabeads (Thermo Fisher) as a positive control. Following the incubation period, cells were stained with Zombie NIR (Biolegend) for 10 min at room temperature before staining cells for 20 min at 4°C with a cocktail of the following antibodies diluted in PBS (HyClone) 1% BSA (Sigma Aldrich): anti-CD4 BV650, anti-CD8 APC, anti-CD215(IL-15Rα) PE, anti-CD3 Percy5.5. Details of the antibodies are reported in **Supplementary Table 1**. Cells were acquired on a BD LSR Fortessa X20 cytometer.

### Flow cytometry staining and PBMC stimulation

PBMCs were thawed and either stained *ex vivo* or stimulated in AIMV 2% FCS with or without peptide pools from SARS-CoV-2 spike, membrane (M), nucleocapsid (N), HCMV pp65 (all 1 μg/ml), or with PMA/ionomycin (PMA 10 ng/ml, ionomycin 100 ng/ml, Sigma Aldrich) for 5 h at 37°C in the presence of brefeldin A (BD, 5 μg/ml). To assess degranulation, CD107a antibody was added to the cells at the beginning of the stimulation. Cells were stained with a viability dye Zombie Aqua (Biolegend) for 10 min at room temperature and then with antibodies targeting surface markers (20 min 4°C, diluted in PBS 1% BSA (Sigma Aldrich)). Cells were fixed overnight in eBioscience Foxp3/Transcription factor fixation/permeabilization buffer (Invitrogen), and intracellular staining was performed for detection of Ki67, granzyme B or intracellular cytokines (30 min 4°C). Cells were acquired on a BD LSR Fortessa X20 and data analysed using FlowJo software v10.8.1. A complete list of antibodies is included in **Supplementary Table 1**.

### Flow cytometry data analysis

Flow cytometry data was analysed in parallel by manual gating methods and by using unsupervised multi-dimensional algorithms. For the latter analysis we concatenated the flow cytometry standard (FCS) files containing the data from 56 patient samples and performed a FlowSOM clustering analysis and visualized identified clusters using uniform manifold approximation and projection (UMAP) for different cell populations (e.g., CD3^+^, CD4^+^ and CD8^+^). FCS files from 56 patients were concatenated after downsampling and UMAP and FlowSOM analyses were done by using a plugin in FlowJo version 10.8.1. All FCS files for the experiments included in this manuscript can be accessed here on FlowRepository: https://flowrepository.org/id/RvFrH1EpMcUjkbPblOQEGqLxzP6yoJ9saVNwr3QODgh3fjQAvWrLnnALOrxkJ4V7

https://flowrepository.org/id/RvFrqLgRoGU9xibsHtTgV8rfoTLT7yODl6RwucTLXpe12fedxUkv8xu1AOnbhqr6

https://flowrepository.org/id/RvFrB4eRWVJ6jcrkYmv024s1VsMlCJTDAxddvgufBqhOBl8zci2FPXMhjN6u1kwl

https://flowrepository.org/id/RvFrXQQJgmxoVKhnxCyftFv1DLKnUG2zA1Xd0Z1PTOwGkJUTMkNfWGO5zR4q5ncr

### Synthetic peptides

15-mer peptides overlapping by ten amino acid residues and spanning the SARS-CoV-2 spike protein and HCMV pp65 (AD169 strain) proteins were purchased from Mimotopes (Australia). The purity of the peptides was > 80% or > 75%, respectively. Peptides were dissolved as described previously^42^. SARS-CoV-2 membrane and nucleocapsid peptivator peptide pools were purchased from Miltenyi Biotec.

### Enzyme-linked immune absorbent spot assay (ELISpot)

Human IFN-γ ELISpot assays were performed using a human IFN-γ ELISpot BASIC kit (Mabtech). MSIP4W10 PVDF plates (Millipore) were prepared by pre-coating wells in 35% ethanol for 30 seconds and washing them thoroughly with sterile water to remove any residual ethanol. Subsequently the plates were coated with capture antibody (mAb-1-D1K; 15 μg/mL) diluted in PBS and incubated overnight at 4°C (Sigma Aldrich). Cryopreserved PBMC were thawed and rested at 37°C, 5% CO_2_ for 5-6 hours. Coated plates were washed 5 times in sterile PBS and blocked for 1-2 hours using R10 medium which is composed of 0.2 µm filtered RPMI 1640 medium supplemented with 10% FCS, 2 mM glutamine, penicillin (100 units/ml) and streptomycin (100 μg/ml). 2 x 10^5^ PBMCs were added to each 96 well plate with or without peptide pools (as indicated) in a total volume of 100 µl in R10. PBMCs incubated with R10 medium alone were used as negative (unstimulated) controls. Peptide pools spanning spike (S1, S2, S3 and S4), nucleocapsid (N1 and N2), membrane protein (M) and HCMV pp65 were used at a final concentration of 2 µg/ml. PBMC stimulated with PMA at 1 μg/mL and ionomycin at 10 μg/mL (Sigma Aldrich) as a positive control. All conditions (positive and negative controls and peptide stimulated wells) were performed in duplicate. Plates were incubated for 16-18 hours at 37°C, 5% CO_2_ and developed as per manufacturer’s instructions. Developed plates were protected from light and dried for 24-48 hours before image acquisition using C.T.L. ImmunoSpot S6 Ultra-V Analyzer. All plates were read using the same settings. Spot forming units (SFU) for each peptide pool were calculated after subtraction of average background calculated from negative control wells. Negative values after background subtraction were adjusted to zero. Responses were considered antigen-specific when spot counts after background subtraction of relevant peptide pool exceeded 2x standard deviations of the unstimulated control wells, as described.^43^

### Cytokine analysis

A customized Luminex assay was used to measure the following 23 analytes (cytokines/chemokines) in patients’ plasma samples: granulocyte macrophage colony stimulating factor (GM-CSF), interferon alpha (IFN-α) and gamma (IFN-γ), interleukin-1a (IL-1α), IL-1β, IL-2, IL-4, IL-5, IL-6, IL-7, IL-8 (CXCL8), IL-10, IL-12p70, IL-13, IL-15, IL-17A (CTLA-8), IL-18, Interferon gamma-induced protein 10 (IP-10 or CXCL10), Monocyte Chemoattractant Protein-1 (MCP-1 or CCL2), Monocyte chemoattractant protein-4 (MCP-4), Macrophage inflammatory protein (MIP-1α), MIP-1β, and Tumor Necrosis Factor-alpha (TNF-α). The ProcartaPlex Multiplex Immunoassay kit (Thermo Fisher Scientific, PPX-23-MXWCXFA) was used according to the manufacturer’s recommendations. Samples were acquired on Luminex200 Analyser Instrument using the xPONENT basic plus partner analytics. Data were analyzed using Invitrogen ProcartaPlex Analysis App on the Thermo Fisher Connect™ Platform. The 2 plasma samples from the acute time points that were RT-PCR positive for SARS-CoV-2 were heat inactivated for 30 min at 56°C prior to their use in the Luminex assay. For the matching 3-12 month plasma samples, Luminex was performed in parallel on samples that had either been heat inactivated or not, and measurements of all detectable analytes were comparable in these two conditions. Only the data from the non-heat inactivated 3-12 months samples was included in Figure 3. Measurement for each sample is performed in duplicates. The following outliers which exceeded by 100 times the minimal value for the corresponding cytokines in all patients were excluded from analysis: for 2 patients (severe disease) we excluded data points for GM-CSF, IFN-g, IL-1α, IL-1b, IL-2, IL-4, IL-6, IL12p70, IL-13, IL-17A, TNF-a at 3 and/or 12 months; for one of these patients we also excluded MIP1-b at 3 months; GM-CSF was excluded for an additional patient (moderate disease) at 12 months.

### Spike-RBD Antibody Bridging LIPS assay

This assay was performed as described previously^44^. Briefly, spike antigen diluted to 2.5 ng/µl in 40 µl PBS was pipetted into every well of a 96-well high-binding OptiPlate™ (Perkin-Elmer, Waltham, MA, USA) and incubated for 18 hrs at 4°C. The plate was washed 4 times with 20 mM Tris 150 mM NaCl pH 7.4 with 0.15% v/v Tween-20 (TBST) and blocked with 1% Casein in PBS (Thermo Scientific, Waltham, MA, USA). The plate was left to air-dry for 2-3 hrs before being stored with a sachet of desiccant in a sealed plastic bag at 4°C and used within three weeks. The Nluc-RBD antigen was diluted in TBST to 10x10^6^+/-5% LU per 25 µl. Sera (1.5 µl, 2 replicates) were pipetted into a 96-well plate and incubated with 37.5 µl diluted labelled antigen for 2 hrs. Of this mixture, 26 µl was transferred into the coated OptiPlate and incubated shaking (∼700 rpm) for 1.5 hrs. The plate was washed 8 times with TBST, excess buffer was removed by aspiration, then 40 µl of a 1:1 dilution of Nano-Glo® substrate (Promega) and TBST was injected into each well before counting in a Hidex Sense Beta Luminometer (Turku, Finland). Units were interpolated from LU through a standard curve.

### Statistical analysis

Statistical analysis was performed using either GraphPad Prism v9.4 (GraphPad Software, San Diego, CA) and R version 4.0.4. The statistical tests and post-hoc corrections for multiple testing used are indicated in each figure legend. PP-values are indicated as follows: *p < 0.05, **p≤0.01; ***p≤0.001. For comparison of the immune data and long COVID symptoms, Poisson regression was used with number of symptoms at 3 months as the outcome variable, and each individual cellular marker/cytokine as the explanatory variable. Analyses were performed unadjusted and adjusted for age and sex. Due to the number of comparisons, FDR correction was performed at a value of 5%. Analysis was performed using the tidyverse package. The script and data are publicly available here: https://github.com/gushamilton/discover_long_covid

## Supplementary data legends

**Supplementary Figure 1.**
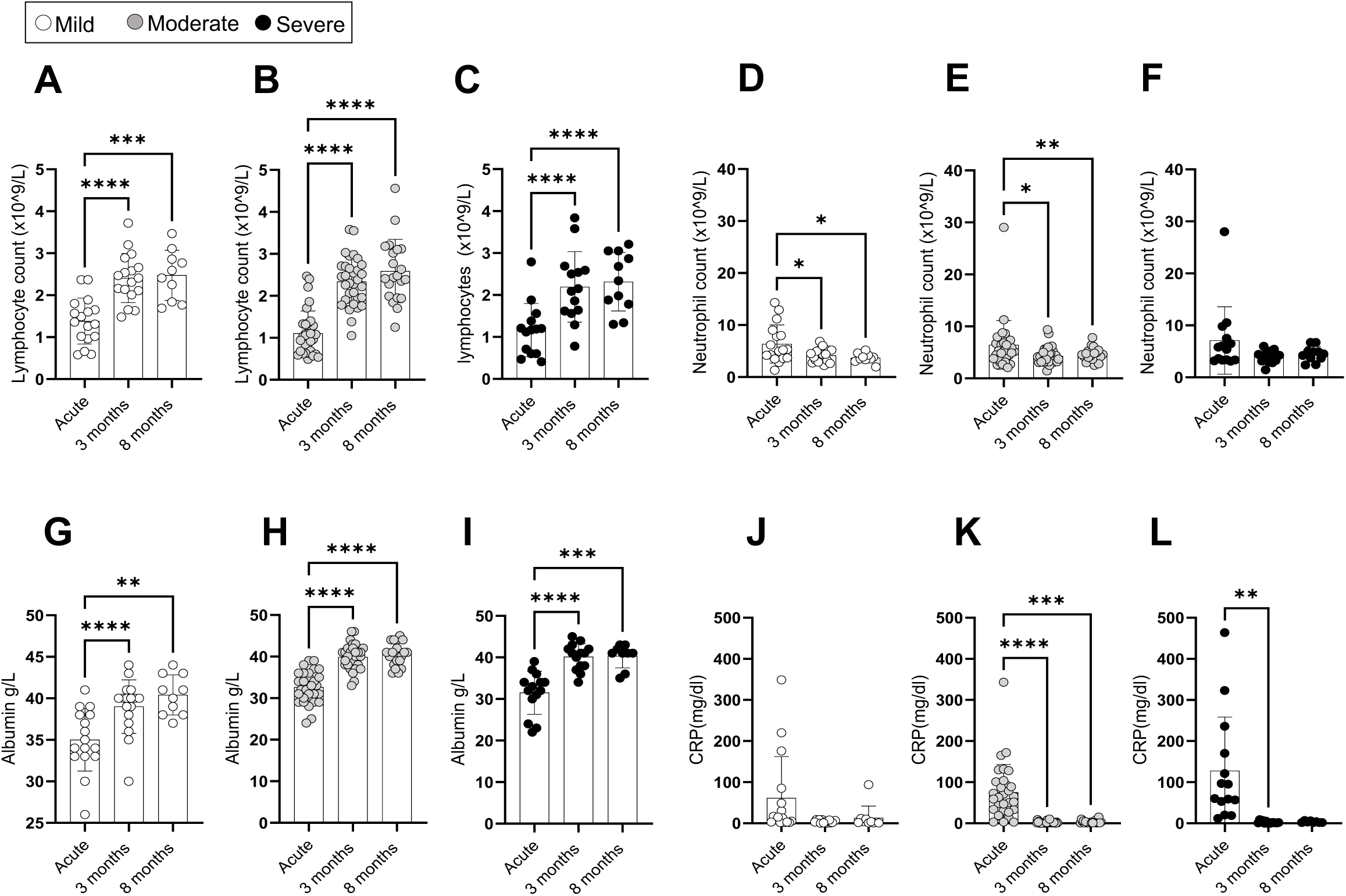
Dynamic changes of immune populations and inflammatory markers in COVID-19 patients at acute illness, 3- and 8-months post admission. A-F. Lymphocyte (A-C) and neutrophil (D-F) counts during acute illness, at 3 and 8 months post admission in patients with mild (A, D), moderate (B, E) and severe (C, F) disease. G-L. Albumin (G-I) and CRP (J-L) levels during acute illness, at 3 and 8 months post admission in patients with mild (G, J), moderate (H, K) and severe (I, L) disease. Data from mild, moderate and severe patients are indicated with white, grey and black symbols; Data are shown as a mean +/- SD; * p < 0.05, ** p≤0.01; *** p≤0.001; Statistics were calculated by One-way Anova, with Geisser-Greenhouse correction for multiple testing.

**Supplementary Figure 2.**
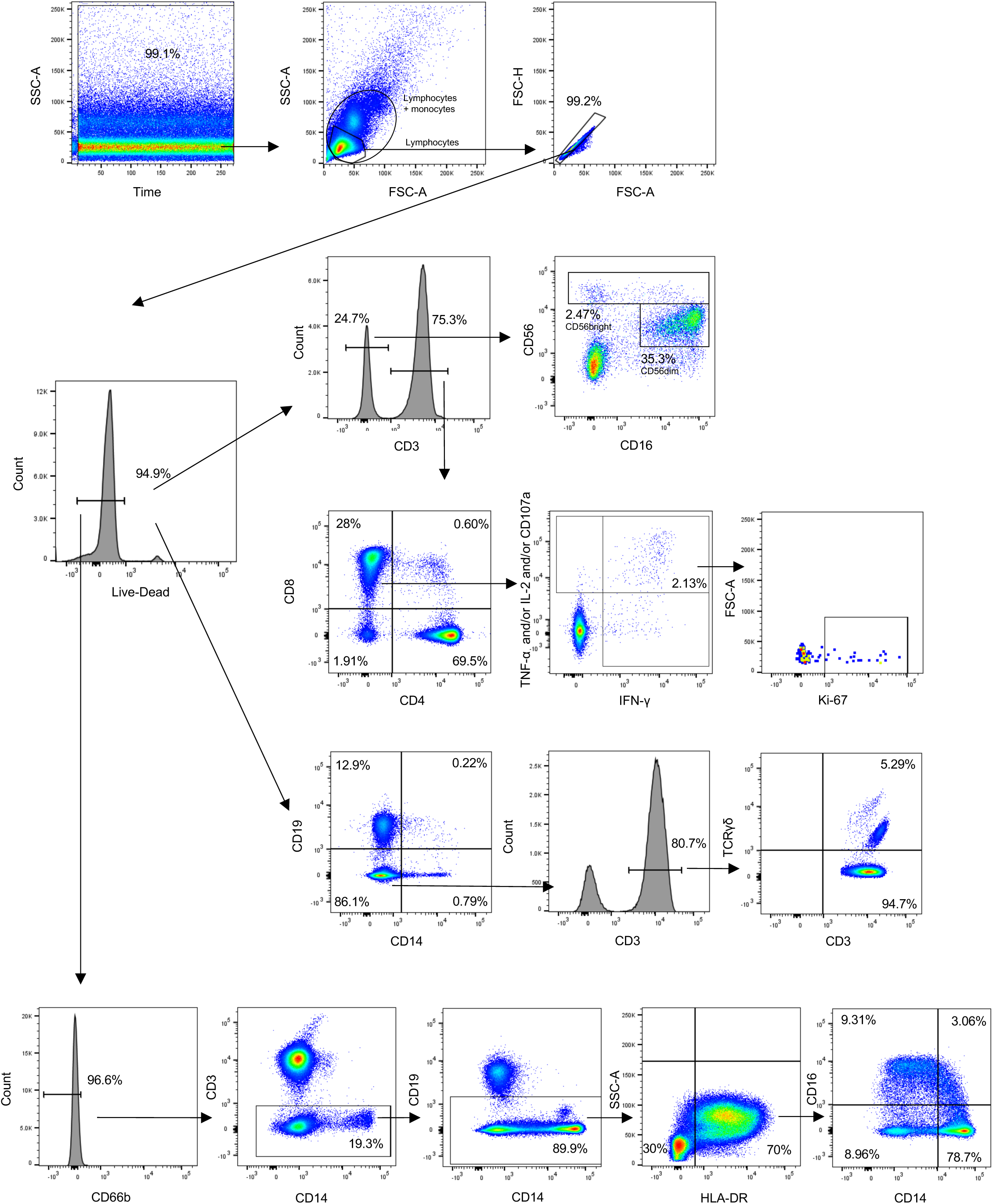
Gating strategy used to identify CD4^+^, CD8^+^ and TCR-γδ T cells, NK cells and monocytes.

**Supplementary Figure 3.**
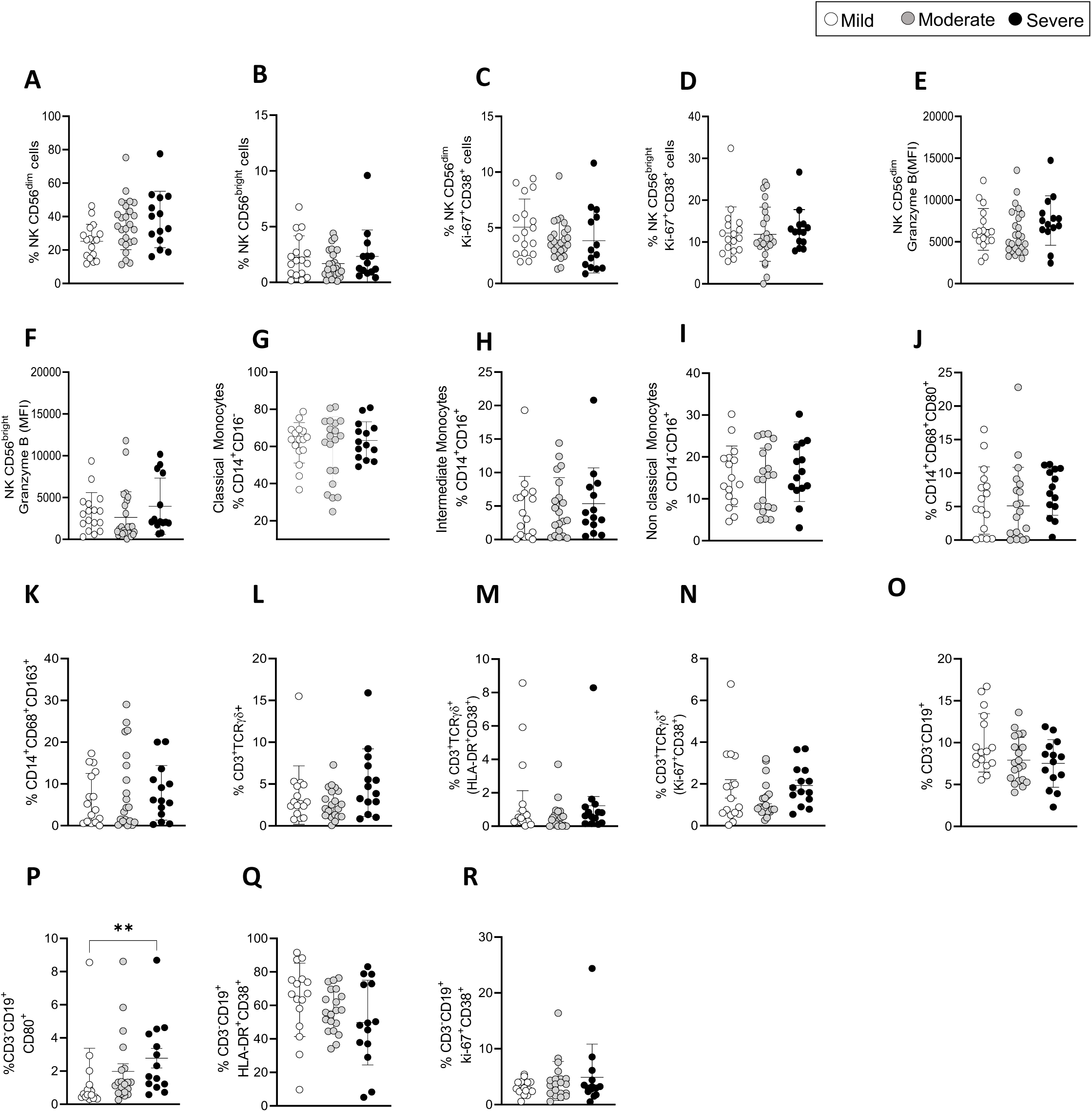
Immune cell populations in COVID-19 patients at 3 months post admission. A-F. Shown are the percentages of CD56^dim^ (A), CD56^bright^ NK cells (B), Ki67+CD38+ CD56^dim^ (C) and Ki67+CD38+ CD56^bright^ NK cells (D). Granzyme B expression on CD56^dim^ (E) and CD56^bright^ (F) NK cells is shown as Mean Florescence Intensity (MFI). G-K. Shown are the percentages of classical (G: CD14+ CD16), intermediate (H: CD14+CD16+) and non-classical (I: CD14-CD16+) monocytes, activated CD14+ CD80+CD86+ (J) and CD14+CD86+CD163+ cells (K). L-N Frequencies of TCR-γδ T cells (L), activated HLA-DR+CD38+ (M) and activated/proliferating Ki67+CD38+ (N). Shown are the percentages of CD3-CD19+ B cells (O), activated B cells CD80+ (P) and HLA-DR+CD38+ B cells (Q) and responding Ki67+CD38+ B cells (R). O-R. Data are visualized as mean +/- SD; Statistics were by calculated One-way ANOVA test (Kruskal-Wallis test) with Dunn’s correction for multiple testing.

**Supplementary Figure 4.**
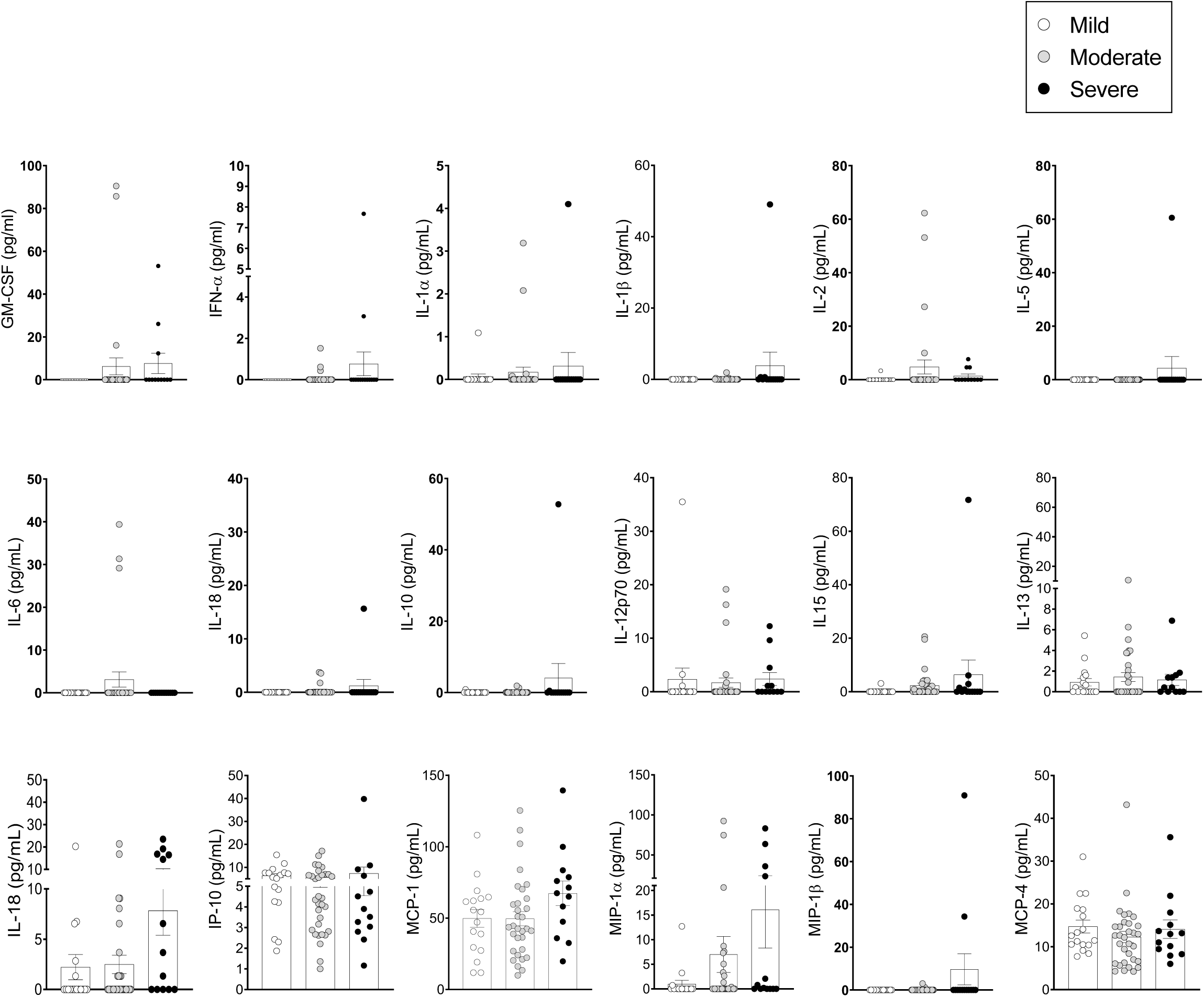
Pro-inflammatory cytokines/chemokines in the plasma of COVID-19 patients at 12 months post admission. Cytokines/chemokines were measured in the plasma of patients by Luminex. A. Analyte levels (pg/ml) are shown for mild, moderate and severe patients at 3 months post-admission (63 samples: Mild: N=17; Moderate: N=32; Severe: N=14, depicted in white, grey and black symbols, respectively). Data are visualized as mean +/- SEM. No statistically significant differences were detected for the cytokines included here, as calculated by One-Way ANOVA test (Kruskal-Wallis test) with Dunn’s correction for multiple testing.

**Supplementary Table 1.**
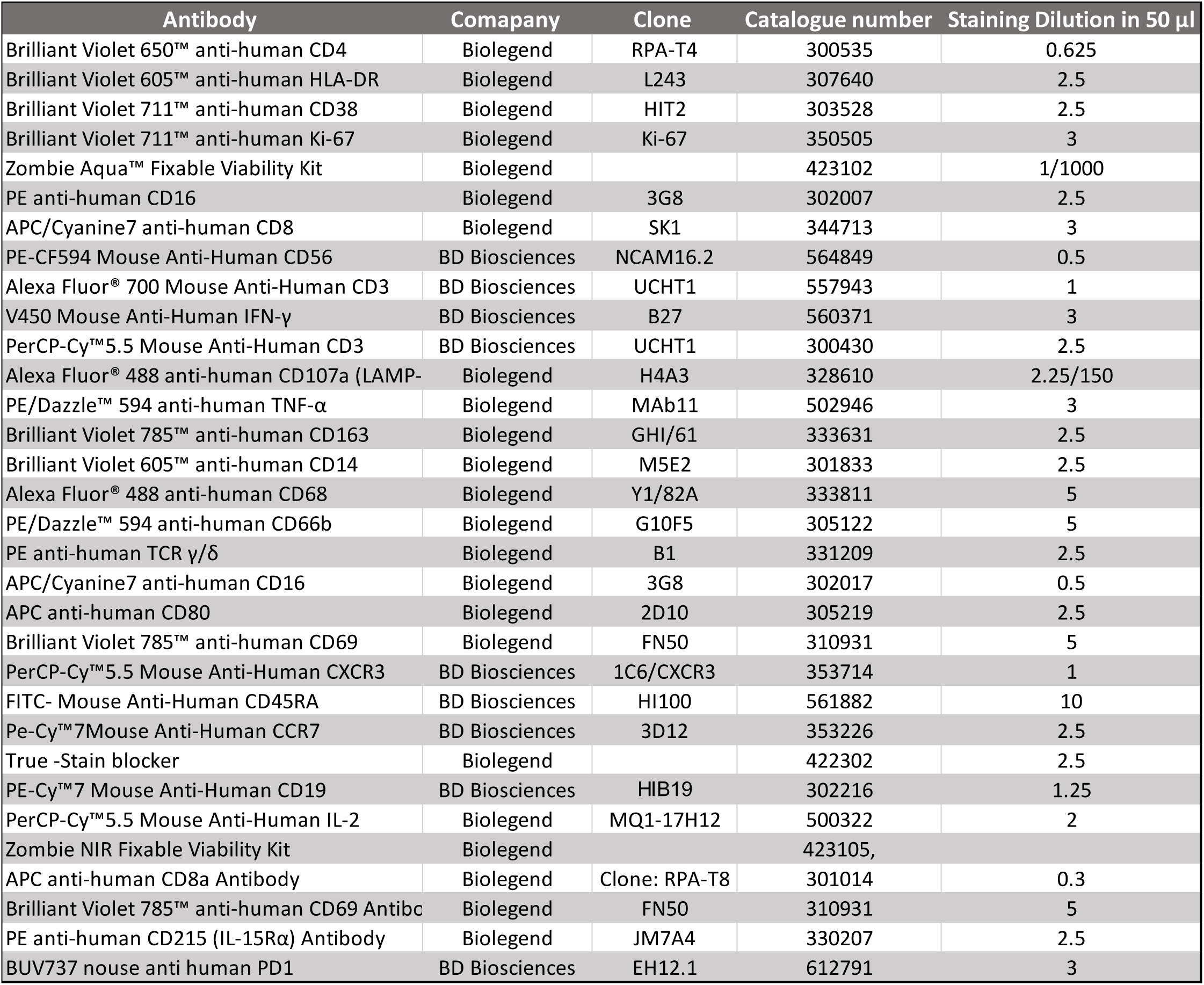
Details of the antibodies used for flow cytometry.

**Supplementary Table 2.**
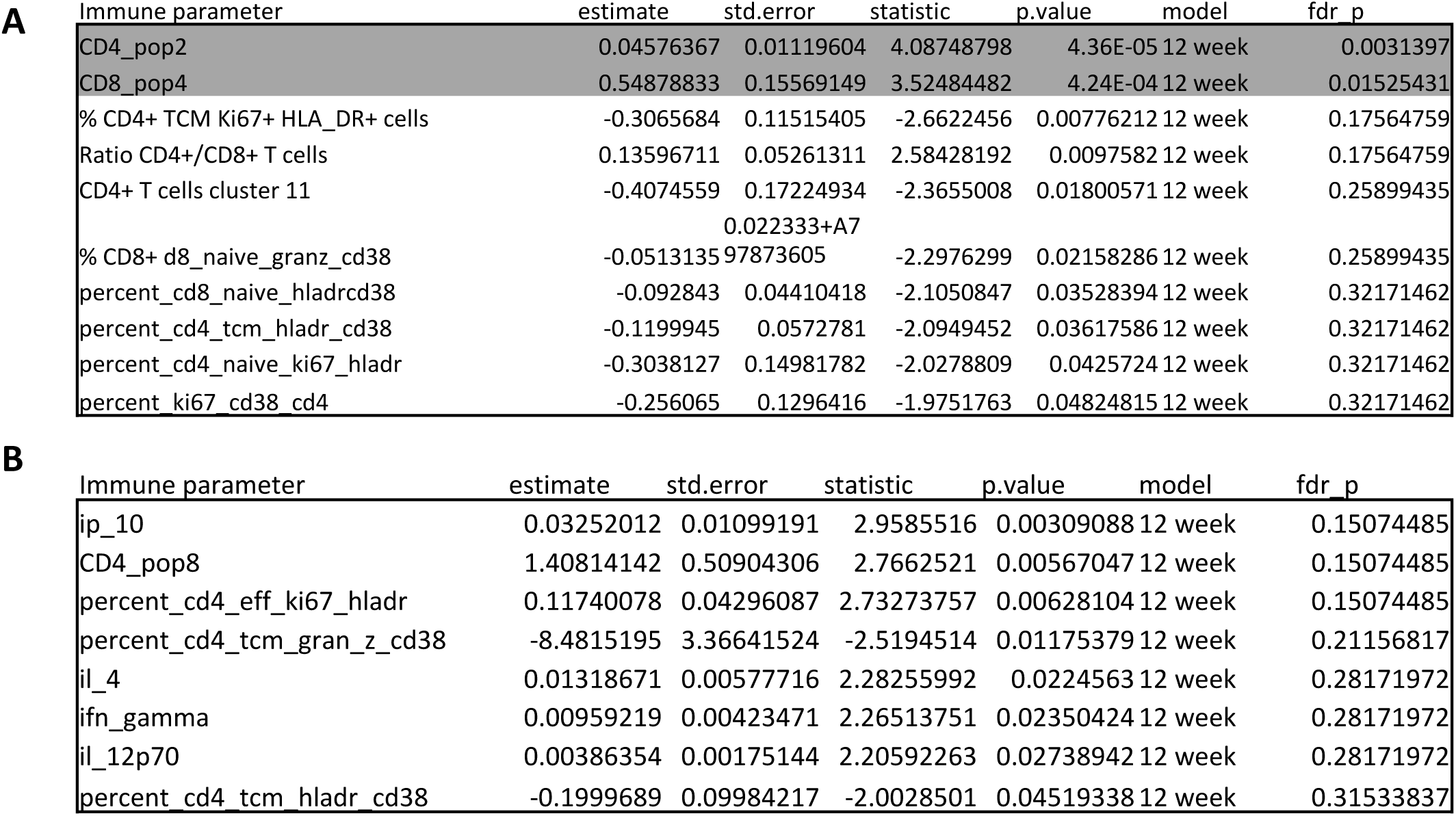
A, B. Associations between immune parameters and symptoms at 3 months in either unadjusted (A) or adjusted (B) Poisson regression models. P values are shown with or without FDR correction (fdr_p or p values, respectively) for both adjusted and unadjusted models. Highlighted in grey are the 2 immune parameters that significantly correlated with symptoms after FDR correction. Only parameters with FDR uncorrected p-values <0.05 are included in the tables.

